# Safety and efficacy of the two doses conjugated protein-based SOBERANA-02 COVID-19 vaccine and of a heterologous three-dose combination with SOBERANA-PLUS: double-blind, randomised, placebo-controlled phase 3 clinical trial

**DOI:** 10.1101/2021.10.31.21265703

**Authors:** María Eugenia Toledo-Romaní, Mayra García-Carmenate, Carmen Valenzuela-Silva, Waldemar Baldoquín-Rodríguez, Marisel Martínez-Pérez, Meiby Rodríguez-González, Beatriz Paredes-Moreno, Ivis Mendoza-Hernández, Raúl González-Mujica, Oscar Samón-Tabio, Pablo Velazco-Villares, Juan Pablo Bacallao-Castillo, Ernesto Licea-Martín, Misladys Rodríguez-Ortega, Nuris Herrera-Marrero, Esperanza Caballero-González, Liudmila Egües-Torres, Reinaldo Duartes-González, Serguey García-Blanco, Suzette Pérez-Cabrera, Santos Huete-Ferreira, Kirenia Idalmis-Cisnero, Omayda Fonte-Galindo, Dania Meliá-Pérez, Ivonne Rojas-Remedios, Delaram Doroud, Mohammad Mehdi Gouya, Alireza Biglari, Patrick Van der Stuyft, Sonsire Fernández-Castillo, Yanet Climent-Ruiz, Yury Valdes-Balbín, Dagmar García-Rivera, Vicente Verez-Bencomo, the SOBERANA Phase 3 team

## Abstract

**Background:** SOBERANA-02 is a COVID-19 conjugate vaccine (recombinant RBD conjugated to tetanus toxoid). Phases 1/2 clinical trials demonstrated high immunogenicity, promoting neutralizing IgG and specific T-cell response. A third heterologous dose of SOBERANA-Plus (RBD-dimer) further increased neutralizing antibodies.

**Methods:** From March 8^th^ to September 30^th^, 2021 we conducted in Havana, Cuba a multicentre randomized, double-blind, placebo-controlled, phase-3 trial evaluating two doses of SOBERANA-02 and a heterologous scheme with one dose SOBERANA-Plus added to it. Participants 19–80 years were randomly assigned to receiving 28 days apart either the two or three dose scheme or placebo. The main endpoint was vaccine efficacy in preventing the occurrence of RT-PCR confirmed symptomatic COVID-19 occurring at least 14 days after the second or third dose in the per-protocol population. We also assessed efficacy against severe disease and, in all participants receiving at least one vaccine/placebo dose, safety for 28 days after each dose.

**Finding:** We included 44·031 participants in a context of Beta VOC predominance, with this variant being gradually replaced by Delta near the trial end. Vaccine efficacy in the heterologous combination was 92·0% (95%CI 80·4–96·7) against symptomatic and 100% against severe COVID-19. Two doses of SOBERANA-02 was 69·7% (95%CI 56·5-78·9) and 74·9% (95%CI 33·7-90·5) efficacious to protect against symptomatic and severe COVID-19, respectively. The occurrence of serious and severe AEs was very rare and equally distributed between placebo and vaccine groups. Solicited AEs were slightly more frequent in the vaccine group but predominantly local and mostly mild and transient.

**Interpretation:** Our results indicate that the straightforward to manufacture SOBERANA vaccines are efficacious in a context of Beta and Delta VOC dominance and that they constitute an attractive, feasible option for low- and middle-income countries, where besides financial constraints ease of vaccine storage and distribution is of concern.

**Funding:** This study received funds from Finlay Vaccine Institute and National Fund for Science and Technology (FONCI-CITMA-Cuba, contract 2020–20). of Ministry of Science, Technology and the Environment (Contract Project-2020-20) in Cuba.

## Introduction

Immunization with an effective vaccine constitutes the main tool to fight the COVID-19 pandemic, and achieving high vaccination coverage is critical to control the emergence and spread of new SARS CoV-2 variants.^1^ Clinical trials provided evidence on the efficacy of vaccines based on different technologies, which received WHO endorsed emergency use authorization.^2^ Notwithstanding, even under optimistic manufacturing and delivery scenarios global equitable access to vaccines is likely to be limited when relying on the currently available products only.^3^

SOBERANA-02 is a subunit conjugated protein-based COVID-19 vaccine. The antigen is the recombinant receptor binding domain (RBD) protein conjugated chemically to tetanus toxoid (TT).^4^ The well-known protein-polysaccharide conjugation technology provided novel, safe and highly immunogenic paediatric vaccines against *Haemophilus influenzae* type b from the 1980s onwards,^5^ and some other for protecting against *Neisseria meningitidis* and *Streptococcus pneumoniae*.^6, 7^

In phase 1 and 2a clinical trials two doses of SOBERANA-02 induced neutralizing antibodies and T-cells with IFNγ secretion, indicative of a Th1 pattern.^8^ A single dose of SOBERANA-Plus (RBD-dimer/Alum) demonstrated booster capacity in convalescent COVID-19 patients who had acquired natural immunity against the virus,^9^ through hybrid immunity.^10^ A third heterologous dose of SOBERANA-Plus after two doses of SOBERANA-02 significantly increased neutralizing anti-RBD IgG titres.^8^ These results were confirmed in a phase 2b clinical trial.^11^

To determine whether the immunogenicity results would translate into differentiated clinical efficacy outcomes we set up a phase 3 trial (IFV/COR/09 number, RPCEC00000354) that evaluates the safety and the efficacy of two immunization regimes: two doses of SOBERANA-02, and the heterologous combination thereof with SOBERANA-Plus as a third dose.

## Methods

### Study design and context

A multicentre adaptive, randomized, placebo-controlled, double-blind phase 3 trial was set up to evaluate the efficacy and safety of vaccination against SARS-CoV-2 with 2 doses of SOBERANA-02 (FINLAY-FR-2) -subsequently referred to as So2- and with a three dose heterologous scheme adding SOBERANA-Plus (FINLAY-FR-1A) to it -hereafter So2P. Participants were recruited from March 8^th^ to March 31^th^, 2021.

The study was conducted in Havana, Cuba. Comprehensive primary health care services with a defined population of responsibility are a key element of the Cuban health care system. They consist of family doctor/nurse practices and policlinics. The former is the systems’ entry point, policlinics provide diagnostic and support services and specialized care. The routine identification of COVID-19 cases is based on first line services, which pinpoint suspects and refer to hospitals. Upon a positive RT-PCR test they are hospitalized until symptom resolution and negative PCR test. Contacts of COVID-19 cases are isolated for 5 days and PCR tested routinely.

The epidemiological context at the start of the trial and during So2 efficacy evaluation coincided with the second epidemic wave in Havana (S4. Figure S1; source: Health Information System, Ministry of Public Health) and predominance of VOC Beta (epidemiological week 16 to 22: 83·7% to 72·5%). During So2P efficacy evaluation the Beta strain was partially replaced by Delta (epidemiological week 22 to 28: 72·5% to 29·2% Beta; 5·9% to 70·8% Delta). The latter was associated with a sharp rise of transmission and a significant increase in the incidence of clinical cases.

### Roles and responsibilities

The trial was sponsored by the Finlay Vaccine Institute. A central Research Ethics Committee was appointed *ad hoc* by the Cuban Ministry of Public Health. It approved the protocol (Clinical Trials IFV/COR/09 number, RPCEC00000354) and the informed consent forms. In the event of a medical problem requiring unmasking, approval by the principal investigator was planned for. All principles of the Helsinki Declaration and the International Council for Harmonization guidelines were adhered to.^12^ Full details on procedures are provided in the protocol.

The National Clinical Trials Coordinating Centre (CENCEC) monitored the trial for protocol adherence and observance of Good Clinical Practice, and oversaw data accuracy. An Independent Safety Data Monitoring Board (ISDMB) continuously supervised safety and conducted interim analysis. The ISDMB had access to the case files, confirmed severe cases of COVID-19 illness, and assessed whether any deaths were SARS CoV-2-related.

### Participants, randomization and blinding

In 8 municipalities of west Havana we set up, in existing health infrastructure, 48 vaccination and clinical sites. Practitioners of the family practices linked to each of these sites organized information campaigns in the community and summoned the population between 19 and 80 years old to participate in the study. The family doctors carried out a first eligibility screening before transferring potential participants to the nearest vaccination site, where formal inclusion took place.

Persons between 19 and 80 years old with no known history of SARS-CoV-2 infection that consented to use reliable anticonception if female and in fertile age, and that provided written informed consent for participation were included. Exclusion criteria were, amongst others, previous receipt of a COVID-19 vaccine, acute febrile illness or infectious disease, pregnancy or breastfeeding, any uncontrolled non communicable diseases, history of severe allergic reactions to any component of the vaccines, and current or planned or history of receiving immunomodulatory drugs. A complete list of eligibility criteria is available in the protocol. At inclusion, a blood sample for asserting rapid serologic evidence of SARSCoV-2 prior asymptomatic infection (Realy Tech IgG/IgM, China) was collected in order to subsequently permit safety and efficacy analyses stratified by prior exposure.

Block randomization into study arms and placebo was done at a 1:1:1 ratio stratified by site and by a priori defined risk strata: 19–64 years without risk of severe COVID-19, 19–64 years with risk of severe disease, and ≥65 years. Participants younger than 65 years of age were categorized as being at risk of severe COVID-19 if they had at least one of the following risk factors: obesity (body mass index ≥35), severe malnutrition (BMI < 18.5), hypertension (according to the Ministry of Public Health guidelines),^13^ chronic kidney disease, ischemic heart disease, diabetes mellitus, chronic obstructive pulmonary disease, asthma (grade 2/3), cancer, HIV or primary/secondary immunodeficiency.

The presentation of the vaccine candidates and placebo were undistinguishable. The participants, the involved health care workers and the investigators were masked to group allocation until the decision was taken to administer or complete the three-dose vaccine schedule.

### Products under evaluation

SOBERANA-02 is a subunit protein-based vaccine. The antigen is the recombinant SARS-CoV-2 RBD (25 μg), chemically conjugated to tetanus toxoid and adsorbed on 500 μg alumina.^4, 8^ SOBERANA-Plus antigen is a dimeric RBD (50 μg) adsorbed on 1250 μg alumina.^14, 15^ The placebo contained SOBERANA-02 ingredients except the active principle. All were stored at 2-8°C and distributed daily to the clinical sites.

### Efficacy assessment

The primary endpoint was Vaccine Efficacy (VE) for preventing occurrence of symptomatic COVID-19 confirmed by a positive SARS-CoV-2 RT-PCR nasopharyngeal swab (RT-PCR), with onset at least 14 days after the last injection in the per-protocol population (PPP). Secondary endpoints were the efficacy for the prevention of severe COVID-19 and death attributed to COVID-19. Agreement with case definition criteria for primary and secondary endpoints was judged blindly by COVID-19 hospital doctors and reviewed by specifically trained medical research team members. The study’s Principal Investigator resolved any discrepancy. COVID-19 symptomatic disease was considered if participants had at least one major symptom or sign (dyspnea, oxygen saturation ≤92%, persistent thoracic pain, neurological disorders, clinical or radiographic evidence of pneumonia) or two minor symptoms (fever, chills, myalgia, headache, sore throat, running nose, diarrhea/vomiting, myalgia, malaise, cough, dysgeusia/anosmia). RT-PCR (QIAGEN, Germany) confirmation was performed by the SARS CoV-2 national reference laboratory at the “Pedro Kourí” Tropical Medical Institute, Havana, Cuba.

Severe COVID-19 was defined by the presence of any of the following: dyspnea; cyanosis, pulmonary infiltration/condensation, oxygen saturation ≤90% or assisted mechanical ventilation, acute respiratory distress syndrome, evidence of septic shock, or admission to intensive care. Death was attributed to COVID-19 if the cause could be attributed to a complication of COVID-19.

### Safety assessment

All adverse events (AE) were recorded in face to face medical consultations on the three consecutive days after receiving each dose of vaccine or placebo and by self-report using a diary of adverse events up to 28 days. AEs were classified as solicited (within 7 days after injection) and unsolicited (within 28 days after injection); local and systemic; grade 1, 2 or 3 (according to the Common Terminology Criteria for Adverse Events, version 5.0),^16^ and causally vaccine related or not (during 28 days) following WHO recommendation.^17^

### Statistical analysis (see also S3-Supplemental Statistical Methods)

The trial was designed to test the hypothesis that the risk of symptomatic COVID-19 is reduced >60% compared to placebo group. The lower limit of the 95% confidence interval was ≥30%, which points to the rejection of the null hypothesis (that the efficacy of either vaccination scheme is ≤30%).^18^ The design required to achieve 90% power (and 2.5% 1-sided type I error) to detect a HR of 0.4 (VE >60% to reduce the risk of symptomatic disease compared to placebo), with a null hypothesis of HR 0.7. VE bounds were derived using a Lan-DeMets O’Brien-Fleming approximation spending function. Two interim analyses (IA) were planned at detection of 53 and 106 symptomatic cases meeting the primary outcome definition.

Vaccine Efficacy was calculated in the per-protocol population (PPP) as percentage reduction in the hazard ratio (HR): VE=100 × (1 –HR) %, in the vaccine groups as compared to the placebo group, with HR estimated from a stratified Cox proportional hazards model, taking into account the defined risk strata. To take the evolution of the epidemiological situation over time into account, the VE for So2P was estimated considering the cases occurring in the same calendar period (14 days after dose 3 in So2P and 42 days after the 2nd dose in placebo).

Safety was assessed in all participants in the safety population (those who received at least one injection). Descriptive summary statistics (numbers and percentages) for participants with any solicited AE, unsolicited AE, unsolicited severe AE, serious AE, and AE leading to discontinuation from the trial are provided by arm and after each dose.

A criterion for stopping due to unacceptable toxicity (frequency of serious vaccine related adverse event >1%) was evaluated iteratively with a Bayesian algorithm (see study protocol).

### Decision to stop the trial

Considering the advantageous results demonstrated in the interim analyses and the explosive epidemiological situation, it was deemed unethical to withhold the benefits of the vaccine from individuals in the placebo group. The Central Research Ethics Committee of the Ministry of Public Health compelled the sponsor to start offering to participants randomised in that group the three-dose schedule. As a consequence, the observation period for efficacy estimation of So2P versus placebo was limited in time. Concurrently, So2 participants were invited to receive a third heterologous dose of SOBERANA-Plus.

## Results

### Trial Population

Between March 8–31^th^, 2021, 45.184 volunteer were screened and 44.031 were randomly assigned to So2 (14.679), So2P (14.677) or placebo (14.675) (Figure 1). Participants’ characteristics were balanced between the study arms (Table 1) and the gender and racial distribution were representative of the Cuban demographic structure (see also S4-Table S3 for characteristics by age and risk of severe COVID-19). At baseline, serologic evidence of a previous SARS-CoV-2 infection was detected in 0·3% of participants. All participants received the first dose of the treatment they were allocated to.

**Table 1.**
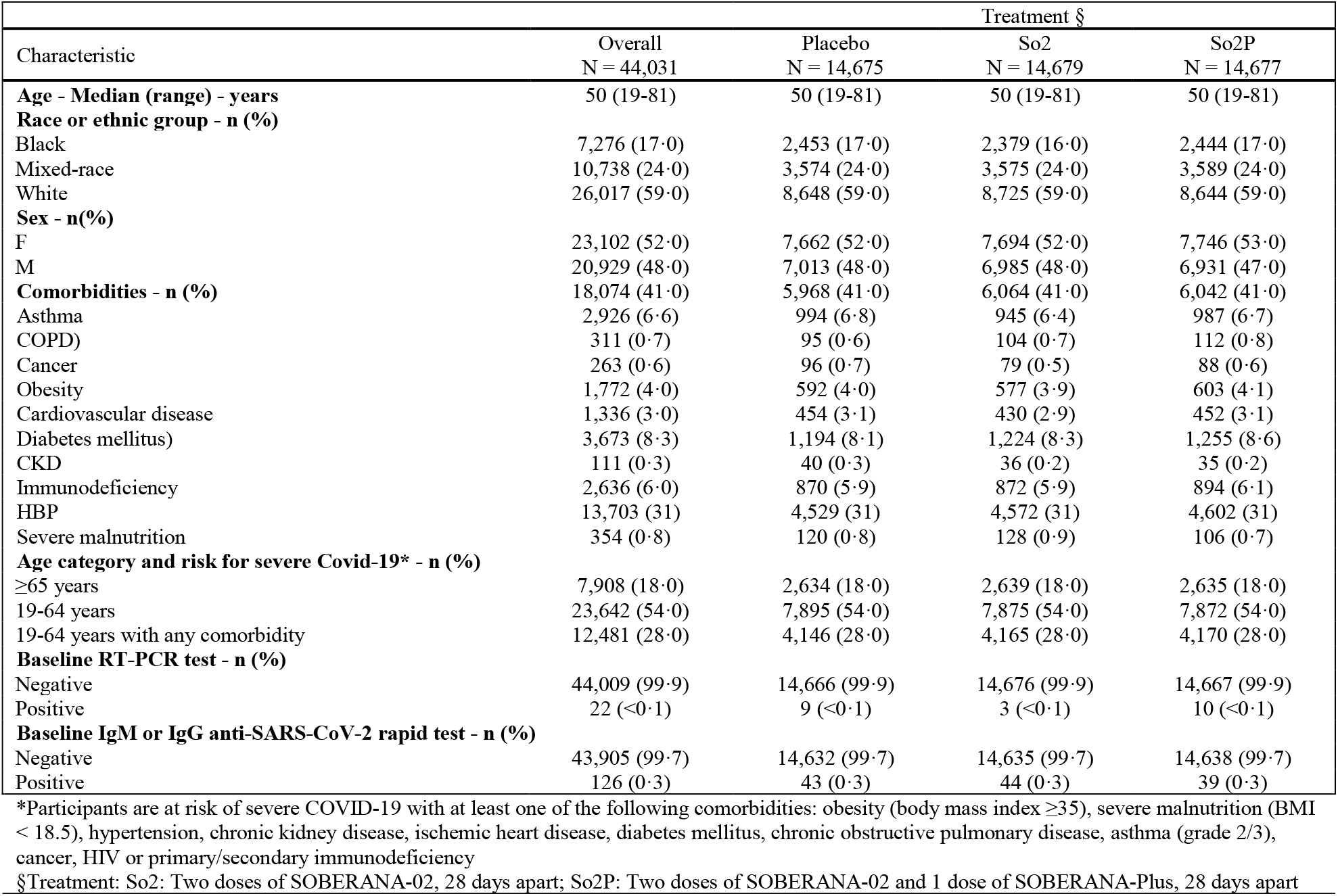
Baseline Demographic and Clinical Characteristics of the randomized participants.

**Figure 1:**
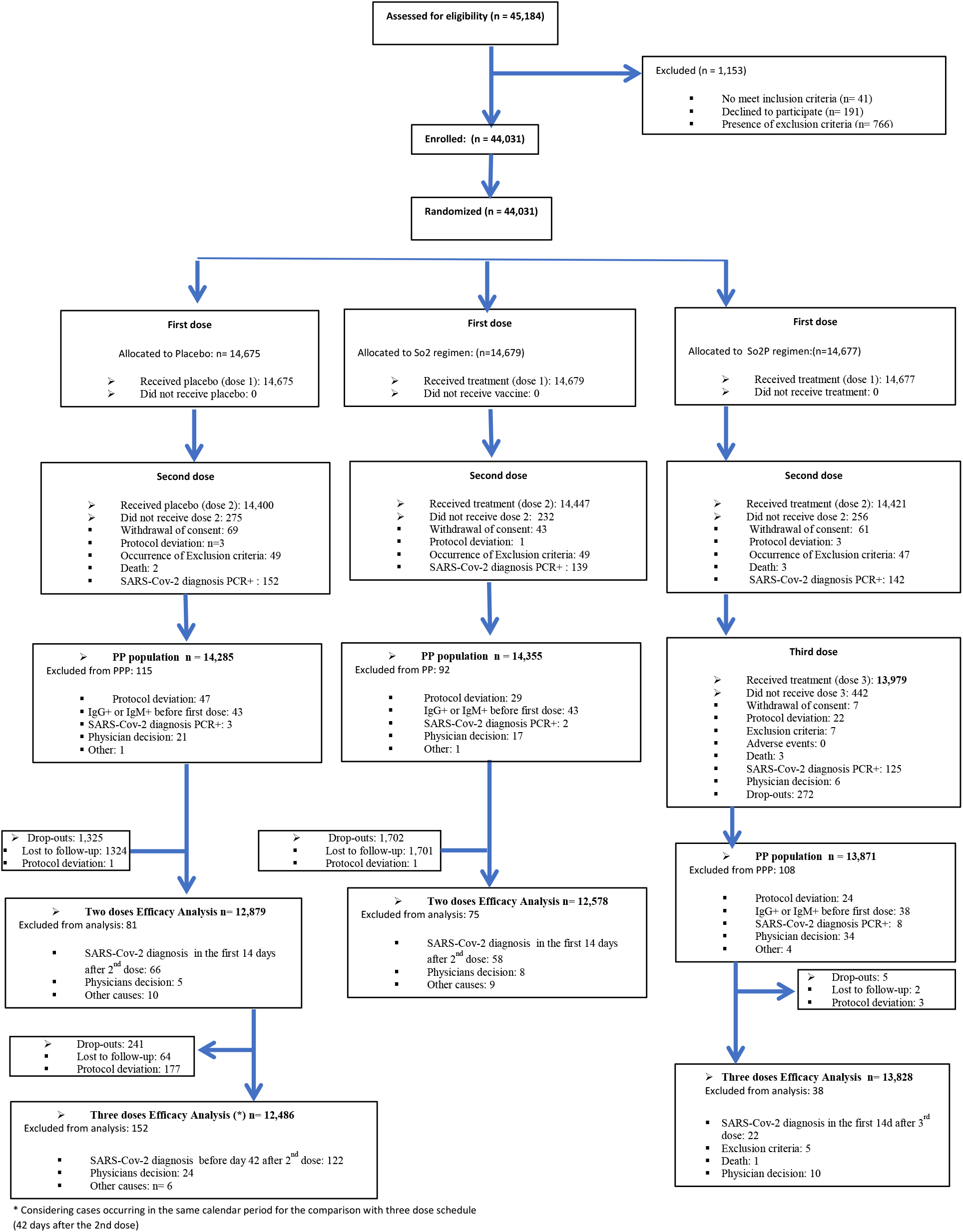
Flow chart. So2 Regimen: two doses of SOBERANA-02 every 28 days; So2P regimen: heterologous combination of two doses of SOBERANA-02 and a third dose with SOBERANA-Plus, every 28 days.

After receiving the first dose, overall 173 (0·39%) participants withdrew their consent and did not receive the second dose and after that dose, 7 (0·05%) from the So2P group withdrew their consent. 433 (0·98%) and 125 (0·85%) participants diagnosed with symptomatic PCR positive SARS-CoV-2 infection were excluded before receiving the second and the third dose, respectively. The PPP primary efficacy analysis included 42.511 subjects receiving the first injection: 14.355 in the So2, 13.871 in the So2P, and 14.285 in the placebo group. Follow-up after the second dose in the two doses analysis totalled 71.202 and 65.136 persons-weeks in the placebo and So2 arm, respectively, while in the three doses analysis it was 20.045 persons-weeks in the placebo and 47.088 persons-weeks in the So2P group.

### Safety

Local solicited Vaccine-Associated Adverse Events (VAAEs) occurred somewhat more frequently in the vaccine group than in the placebo group after the first dose (7·7%, vs. 2·6%, 95%CI for difference: 4·7-5·5) and the second dose (1·9%, vs. 0·4%, 95%CI for difference: 1·3-1·7) (Figure 2, S6-Table S4, S7-Table S5). After the third dose, the frequency of subjects with local AEs dropped to 0·3% (S7-Table S6). In the vaccine groups 97·5% of local adverse events were grade 1 and less than 0·2% grade 3. They lasted a median of 2 days after the first and second dose (25-75^th^ percentile: 1-3 after the first and 1-2 after the 2^nd^ dose), and 2 days after the third dose (25-75^th^ percentile: 0-3). The most common local AE was pain at the injection site (2·6%, 7·5% and 8·7% in placebo and vaccine groups, respectively). Onset of injection-site reactions on or after day 8 were noted in 21 participants (0·05%) after the first dose (4 for placebo and 17 for vaccine groups), in 4 (0·01%) after the second dose (2 for placebo and 2 for vaccine groups) and in 1 participant (0·01%) after the third dose. Local reactions were characterized by erythema, induration, warmth, swelling and pain, and resolved after a median of 1 day (25-75^th^ percentiles: 0·5-2).

**Figure 2.**
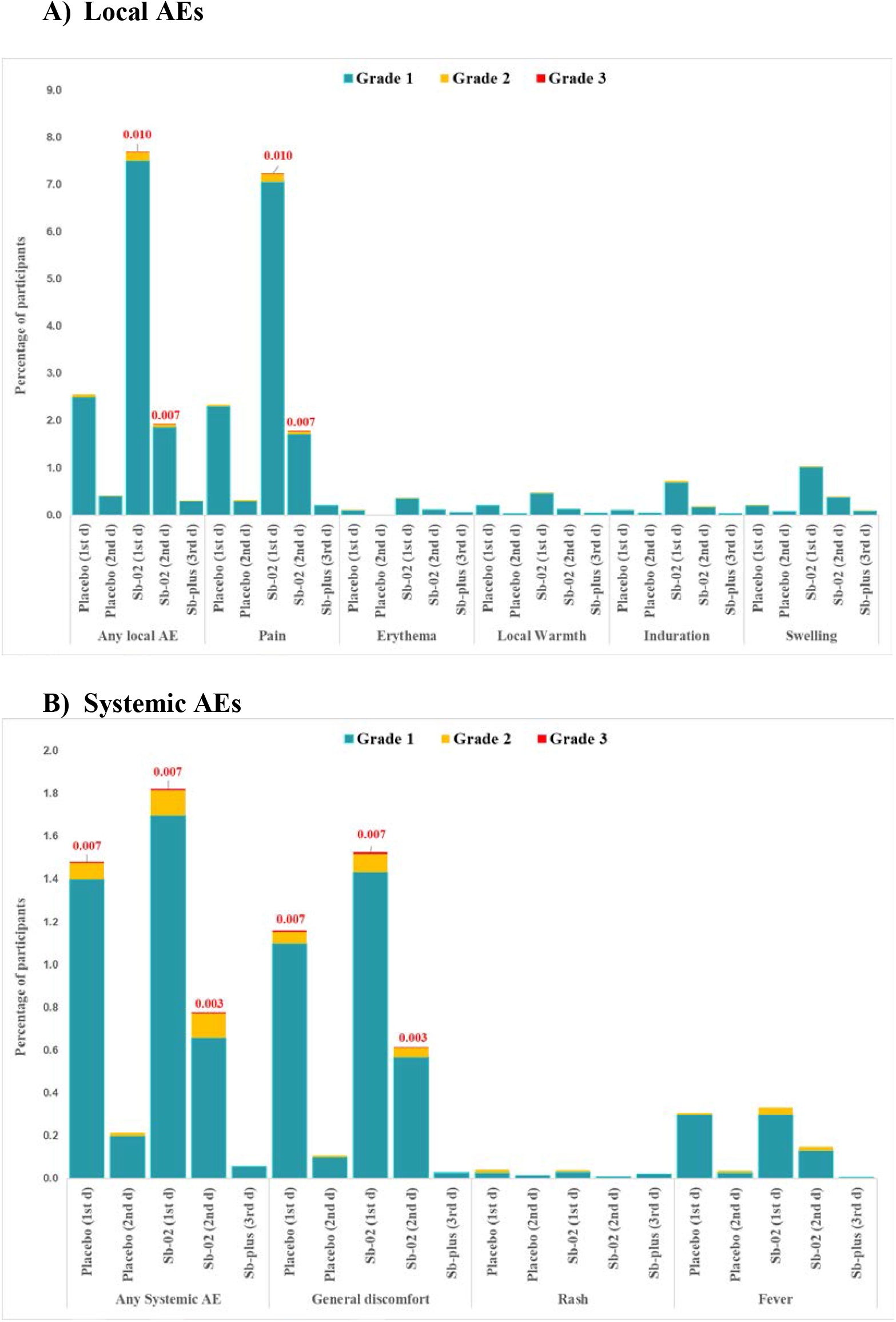
Solicited Vaccine-Associated Adverse Events (VAAE) Within 7 Days After each dose by Grade. Numbers in red above the bars indicate the % grade 3 AE

Solicited systemic VAAEs occurred a little more often in the vaccine groups than in the placebo group after both the first dose (1·8% vs. 1·4%; 95%CI for difference: 0·1-0·6%) and the second dose (0·7% vs. 0·2%; 95%CI for difference: 0·4-0·6%) and dropped to 0·1% after the third dose (Figure 2, S6-Table S4, S7-Table S5, S8-Table S6). Their severity in the vaccine groups changed slightly from 6·5% after the first dose to 8·8% after the second dose for grade 2 events, and from 0·6% to 0·4% for grade 3 ones. Solicited systemic AEs in the vaccine groups lasted a median of 1 day after the first (25-75^th^ percentile: 0-2) and second doses (25-75^th^ percentile: 0-3), and 2 days after the third dose (25-75^th^ percentile: 0·8-7·5). The frequency of solicited AE was very low in subject with serologic evidence of a past SARSCoV-2 infection at intake: 0·1% of local AE after any injection and <0.1% of systemic events.

Unsolicited AE frequency reported during the 28 days following each injection was similar in all groups. The more frequent events (≥1%) were a high blood pressure measurement, mainly in subjects with known hypertension (1·8% for placebo, 2·1% for So2 and 2·2% for So2P) and headache (1·0% for placebo, 1·2% for So2 and 1·5% for So2P) (S9-Table S8). The occurrence of serious (<0·1%) and severe (0·1%) VAAE was equal between groups (S8-Table S7, S10-Table S9). 3·9% of all the AEs were grade 3 in the placebo group against 3·4% and 4·2% for groups So2 and So2P, respectively and 7·9%, 6·7% and 6·7% were serious in these groups, respectively. All-cause mortality was 24 (0·2%) amongst placebo recipients against 9 and 11 (0·1%) in groups So2 and So2P (P=0·014). No death was vaccine related.

### Efficacy

The final analysis of the two-doses schedule included 174 cases of symptomatic COVID-19: 136 in the placebo group (99·9 per 1000 person-years; 95%CI 83·8–118·2) and 38 in the So2 group (30·5 per 1000 person-years; 95%CI 21·6–41·9). This corresponds to 69·7% VE (95% CI 56·5-78·9%; P<0·001) for the prevention of symptomatic SARS-CoV-2 infection by the 2 dose So2 schedule (Table 2, Figure 3, S13-Figure S2). The vaccine efficacy to prevent COVID-19 was congruous across subgroups stratified by demographic and baseline characteristics (S14-Figure S3).

**Table 2.** Vaccine Efficacy to prevent symptomatic COVID-19 disease in the Per-Protocol Population (PPP)

| COVID-19 disease | Two-dose Vaccine Efficacy |  |
| --- | --- | --- |
|  | Placebo | So2 |
| n of events, PPP | 136 | 38 |
| Person- weeks follow-up | 70959 | 64926 |
| Incidence rate per 1,000 Person-Years (95% CI) | 99.9 (83.8; 118.2) | 30.5 (21.6; 41.9) |
| Vaccine Efficacy (95%CI) |  | 69.7 (56.5; 78.9) |
|  | Heterologous three-dose Vaccine Efficacy |  |
|  | Placebo | So2P |
| n of events, PPP | 35 | 6 |
| Person- weeks follow-up | 20047 | 47088 |
| Incidence rate per 1,000 Person-Years (95% CI) | 91.4 (63.4; 126.6) | 6.6 (2.4; 14.5) |
| Vaccine Efficacy (95% CI) |  | 92.0 (80.4; 96.7) |

Vaccine efficacy was defined as 1 minus the hazard ratio (Vaccine vs. placebo) and the 95% confidence interval was estimated with the use of a stratified Cox proportional hazards model

**Figure 3.**
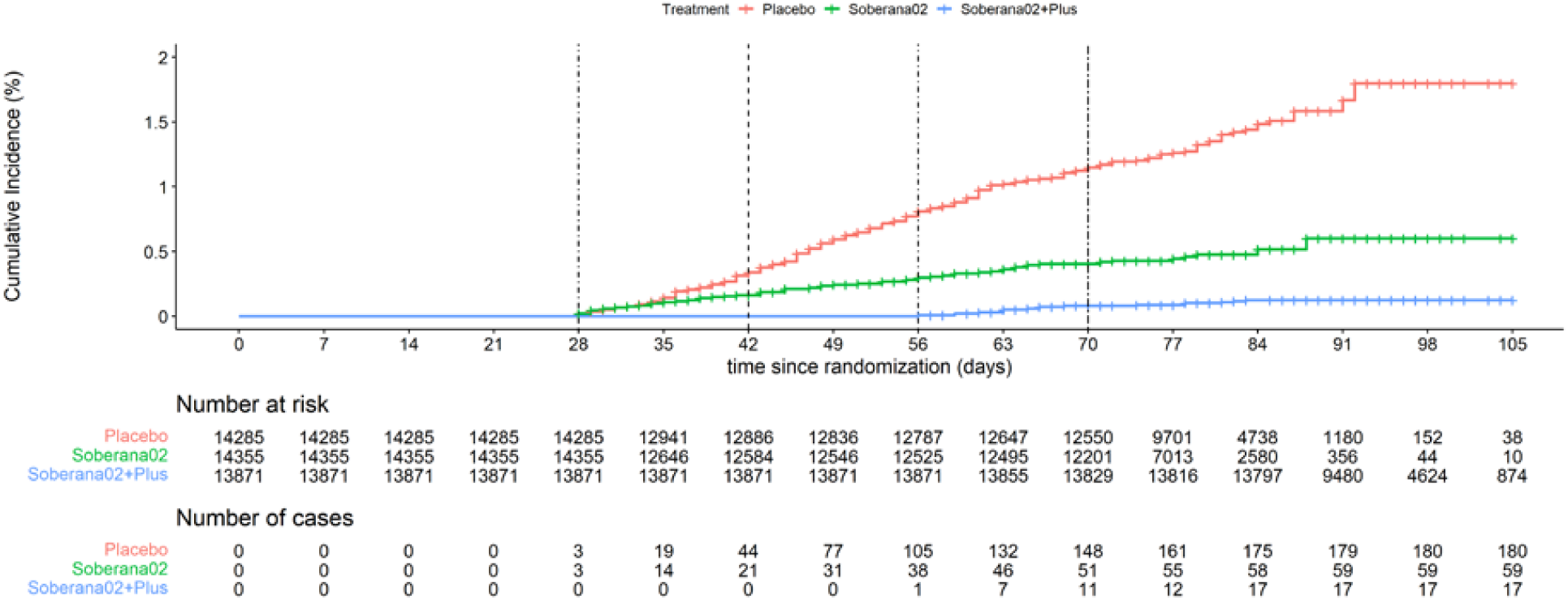
Cumulative incidence of Covid-19 disease. Vertical dashed lines indicate the start of: dose 2 administration, of So2 efficacy evaluation, of dose 3 administration and of So2P efficacy evaluation, respectively

The VE for prevention of severe COVID-19 (23 cases in placebo vs 5 in So2) was 74·9% (95%CI: 33·7-90·5). (S12-Table S10). There were 3 and 2 deaths from COVID-19 disease, respectively; the small number of events precludes a point estimate of VE.

For the final analysis of the three-dose heterologous So2P schedule 41 cases of COVID-19 were included: 35 in the placebo group (91·4 per 1000 person-years; 95%CI 63·4-126·6) and 6 in the So2P group, (6·6 per 1000 person-years; 95%CI 2·4-14·5). This means 92·0% VE (95%CI 80·4–96·7%; P<0·001) for prevention of symptomatic SARS-CoV-2 infection by the 3-dose schedule (Table 2, Figure 3, S15-Figure S4). Subgroups analysis yields remarkably consistent VE estimates, but lacks precision for much of the strata (S16-Figure S5). Severe COVID-19 occurred in 6 and 0 participants in placebo and So2P, respectively, implying 100% VE (S12-Table S10). There were no deaths from COVID-19 disease. Including in the analysis subjects that were PCR negative but had a IgG/IgM positive test at inclusion, indicating past SARSCoV-2 infection, did not affect any of the VE estimates reported in all the above PPP analysis.

## Discussion

The SOBERANA vaccines demonstrated to be efficacious in preventing PCR-confirmed symptomatic SARS-CoV-2 infections among adults aged 19–80 years. Efficacy after two doses SOBERANA-02 administration, 69·7%, was well over the WHO-set 50% threshold.^19^ After completion of the heterologous schedule with SOBERANA-Plus, VE attained an excellent 92·0%. Efficacy to prevent severe COVID-19 was 74·9% and 100·0% for the two dose and heterologous regimen, respectively. The occurrence of serious and severe AEs was very rare and equally distributed between the placebo and vaccine groups. Solicited AEs were slightly more frequent in the vaccine group but predominantly local and mostly mild and transient.

Subunit protein vaccines present clear advantages in terms of manufacturing, storage and distribution.^19^ Still, administration of a third heterologous dose in the So2P regimen, which significantly further increases SOBERANA’s VE, could be construed as a disadvantage compared to COVID-19 vaccines using two dose regimens. However, the need for and benefits of administering a booster dose to the primary schedules of registered vaccines is increasingly recognized.^20, 21^ Furthermore, due to the T-epitopes from the tetanus toxoid carrier protein, SOBERANA-02 -the first conjugate SARS CoV-2 RBD protein based vaccine-induces T-helper response and long lasting immunity.^8, 11^ Heterologous combination for priming and booster using protein vaccines are reported by others with excellent results.^22, 23^

Preclinical results highlighted the stimulation of specific B- and T-memory cells, affinity maturation and a significant and boostable IgG immune response in mice.^4^ The immunogenicity pattern observed in a phase 1 and 2a trials suggested as the best option two doses of 25-μg SOBERANA-02 followed by a third dose of SOBERANA-Plus.^8^ In phase 2b the 4-fold anti-RBD IgG seroconversion rate in 758 volunteers was 76·3 % after two doses and 96·8% after the third heterologous dose, against 7.3% in the placebo group.^11^ Neutralizing IgG antibodies were detected against D614G and VOCs Alpha, Beta, Delta and Omicron, and neutralizing antibodies were still present 7-8 months after the third dose.^11^ The results of the present trial permit to conclude that this leads to improved clinical efficacy of the 3-dose scheme.

A living systematic review reports pooled vaccine efficacy against symptomatic COVID 19 for different platform as: mRNA vaccines 95% (95%CI: 92%-97%); protein subunit vaccines 77% (95%CI 5%-95%); viral vector vaccines 68% (95%CI: 61%-74%) inactivated vaccines 61% (95%CI 52%-68%]. It found that viral vector vaccines decreased overall mortality (risk ratio=0·25; 95%CI 0·09-0·67), but analogous estimates for other platform vaccines are imprecise.^24^ SOBERANA’s performance in the present study compares favourably with the above findings.

Furthermore, the trial was conducted in a challenging epidemiological context: VOC Beta was by far predominant during So2 efficacy evaluation and gradually replaced by VOC Delta after starting So2P evaluation. Beta has demonstrated significant immune evasion ability. It carries the E484K mutation that substantially reduces neutralization by antibodies in sera of convalescent and vaccinated persons.^25^ The few available VE trials on Beta, most of them conducted in South Africa, show efficacy decline with respect to the original SARS-CoV-2 strain. Except for BNT162b2 (Pfizer/BioNTech), with reported efficacy of 100% (95%CI 53·5-100·0),^26^ ChAdOx1-nCoV-19 (AstraZeneca) was not efficacious against mild-to-moderate Beta variant disease,^27^ a single dose Ad26.COV2.S (Janssen) shows 52·0% (95% CI 30·3 to 67·4) against moderate disease,^28^ NVX-CoV2373 (Novavax) was 60·1% (95%CI 19·9-80·1) against symptomatic disease.^29^

Two doses of SOBERANA-02 hence achieved 69·7% (95% CI 56·5-78·9%), quite remarkable efficacy against VOC Beta in our racially diverse Latin-American trial population. One could surmise that, being a conjugated protein-based vaccine that triggers a broad immune response, the efficacy might remain relatively well at par against other immune evading variants. The heterologous combination performed still considerably better. It is, however, not possible to fully unravel this further improved efficacy with a third SOBERANA-Plus dose from the partial replacement of Beta by Delta VOC.

Notwithstanding, both variants have evasion ability and observational studies have been providing evidence of reduced effectiveness of WHO authorised emergency use listing vaccines against both of them.^2^ The encouraging safety profile observed during the phase 2 SOBERANA clinical trial was confirmed in this phase 3 study.^8,11^ Pain at the injection site was not uncommon, akin to what has been observed for other subunit protein and mRNA COVID 19 vaccines,^23^ but mostly mild, while rates of other local reactions were low. Fever and general discomfort were rather uncommon and generally mild, as were other systemic reactions. Serious and severe AEs occurred very rarely and similarly in the vaccine and placebo groups and no death was vaccine related, but sample sizes far beyond those of a typical clinical trial may be needed to adequately appraise these frequencies.

The decision to stop the trial early, remove masking and offer full 3 dose vaccination to all participants constitutes a limitation of our study. It shortened the follow up period for assessing the efficacy of the three dose So2P schedule. However, we still obtained a precise and unbiased VE estimate with narrow CI. Furthermore, to observe waning of protection approximately 6 months follow up would be needed, which can generally only be achieved in observational studies under real life conditions.

Recruitment of participants through family medicine practices, which guaranteed accelerated inclusion reflecting the composition of the population in terms of gender, race and pre-existing conditions, constitutes a strength of this study. Coordination with the health system’s established and well adhered to routine SARS-CoV-2/COVID-19 activities for case ascertainment, hospitalization standardized case management contribute to the strengths.

Overall, our results indicate that the straightforward to manufacture SOBERANA vaccines are efficacious in a context of Beta and Delta VOC dominance and that they constitute an attractive, feasible option for low- and middle-income countries, where besides financial constraints also ease of vaccine storage and distribution is of concern.

## Data Availability

All data produced in the present study are available upon reasonable request to the authors

https://rpcec.sld.cu/en/trials/RPCEC00000354-En

## Data sharing

Protocol (English version) is available at submission. Data about adverse events and VE are shared in the Supplementary Material. Information is also available at the Cuban Public Registry of Clinical Trials, included in WHO International Clinical Trials Registry Platform (https://rpcec.sld.cu/en/trials/RPCEC00000354-En). Other supporting clinical documents, including statistical analysis plan and the informed consent form will be available after publication of this article. Proposals should be sent to. These proposals must be reviewed and approved by the sponsor and the investigator. Finally, a data access agreement must be signed.

## Acknowledgments

We especially thank all the volunteers who participated in the trial and the members of the SOBERANA-Phase 3 clinical trial team (listed in the Supplementary Information) for their dedication and contributions to the trial.

We further thank the IDSMB members: N. A. Jimenez Perez (Chair and Clinical Expert) and C. M. Fonseca Gómez (Clinical Expert) from the Pedro Kourí Tropical Medicine Institute, G. María Suárez Formigo (Immunological Expert) and P. Lorenzo-Luaces Alvarez (Statistical Expert) from the Center for Molecular Immunology, M. Martínez Cabrera (Clinical Expert) from the Ministry of Public Health.

We also thank Rolando Pérez, Ileana Morales and Eduardo Martínez (BioCubaFarma), and Lila Castellanos for their advice and support.

## SUPLEMENTARY INFORMATION

**S1. Table S1.** SOBERANA-2 Phase 3 Team.

| **Process** | **Institution** | **Process team** |
| --- | --- | --- |
| **Project development** | Finlay Vaccine Institute, Laboratory of Synthetic and Biomolecular Chemistry, Center of Molecular Immunology, “Cira Garcia” Clinic, “La Lisa” 1<sup>st</sup> Clinic | D Santana, DG. Rivera, E Ojito, B Sánchez, T Boggiano, R Puga and L Verdecia |
| **Quality management** | Finlay Vaccine Institute | R Martínez, M Naranjo, J Lora, B Guillén, A Santana, T García |
| **Coordination Center** | University of Havana and Finlay Vaccine Institute | Perez C, R Lombana, M Muñoz, Y Rosell, L Fernández, J Torralbas, J Faloh, Y Marrera, O Richards, M Guzmán, A Linen, R Arias, R Varela |
| **Adverse events management** | Havana Municipalities: Playa, Plaza, Habana Vieja, Cerro, Mariano, La Lisa. | L Creagh, B Gómez, Y Cuesta, D Trejo, A Fuentes, I Selman-Housein. |
| **Case management** | Havana Hygiene and Epidemiological Center, University of Computer Science, Public Health Hotel “Linea e I”, “Julio Trigo” Hospital, “Manuel Fajardo Hospital” and National System for Medical Emergencies | N Roca, I Morgado, L Valdés, D Guerra, M Delgado, C Olivera, L López, H Fundora, T. Lok, T Torres, A Campo, Y Olivera-Nodarse |
| **Data management** | Finlay Vaccine Institute, “José Antonio Echevarría” Havana Technological University, UCT Geocuba Research and Consulting and Information’s Technology Company | D Díaz, M Medina, Y Domínguez, J Espi, M Fontanies, M Cárdenas, A Carbajal, G Martínez, C Guach, G Guerrero, S Díaz, D Céspedes, C Hechavarria, D Rodríguez, A Perdomo, D Treto, M Trigo, R Montes, T Blanco, A Suarez, R Marrero H<sup>25</sup>, J Farre, W. Torres, J López, J Vega, A de Céspedes, O Medina, R de la Nuez, A Bravo, O Riquenez, D Echevarría, K Castro. |
| **Product management and masking** | Distributing Medicines Specialized organization (EMCOMED) and Finlay Vaccine Institute | Chacon A, Z Cepero, L Fernández, Y Reina, M Calviño, M González, N Rey, A Pérez, M Lazo, A Villegas, A López, R Torres, J Expósito, E Méndez, M Albuerne, O Perdomo, Y Ricardo, A Albernal, Y Salazar, Y Martínez |
| **Monitoring** | National Clinical Trials Coordinating Center | A Arteaga, Amoroto M, J Álvarez, E García, Y Muñoz, A Vargas, I Brito, P Guerra, Y Lobo, Y Chappi, A Maceo |

**S2. Table S2.**
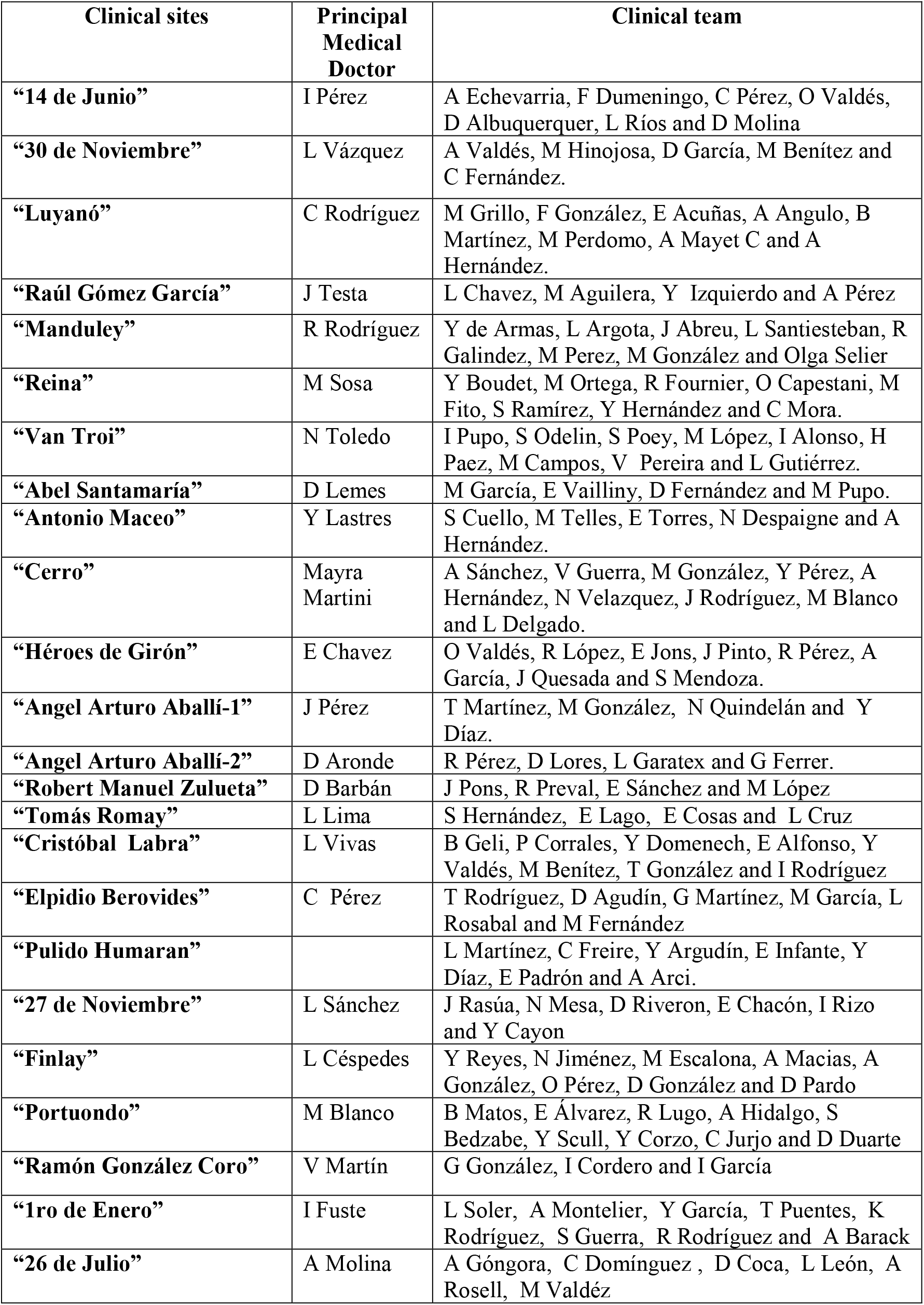
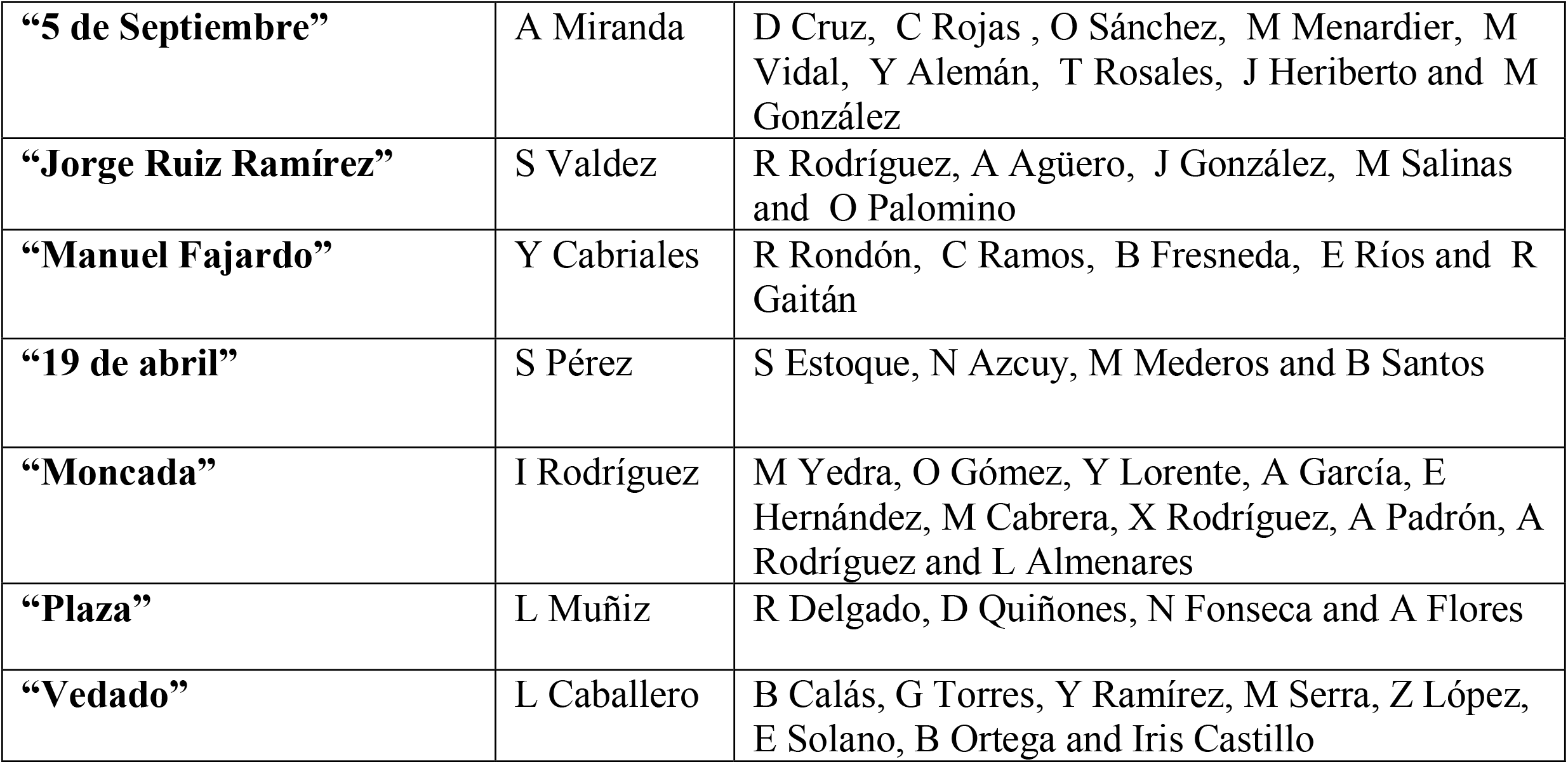
List of SOBERANA-2 Phase 3 Medical Investigators.

### S3. Supplemental Statistical Methods

#### Statistical aspects of the vaccine efficacy analysis

Stratified Cox proportional regression model according to randomization strata was used to evaluate vaccine efficacy (VE), under the assumption of proportional hazard measured as 1 – HR (in each schedule vs. placebo) to test the hypothesis H0: HR ≥ 0.7 (equivalent to VE ≤ 0.3).

Subjects with no confirmed symptomatic illness were censored (assuming that the confirmed illness event did not occur when the subject was under observation). The following intercurrent events were considered:

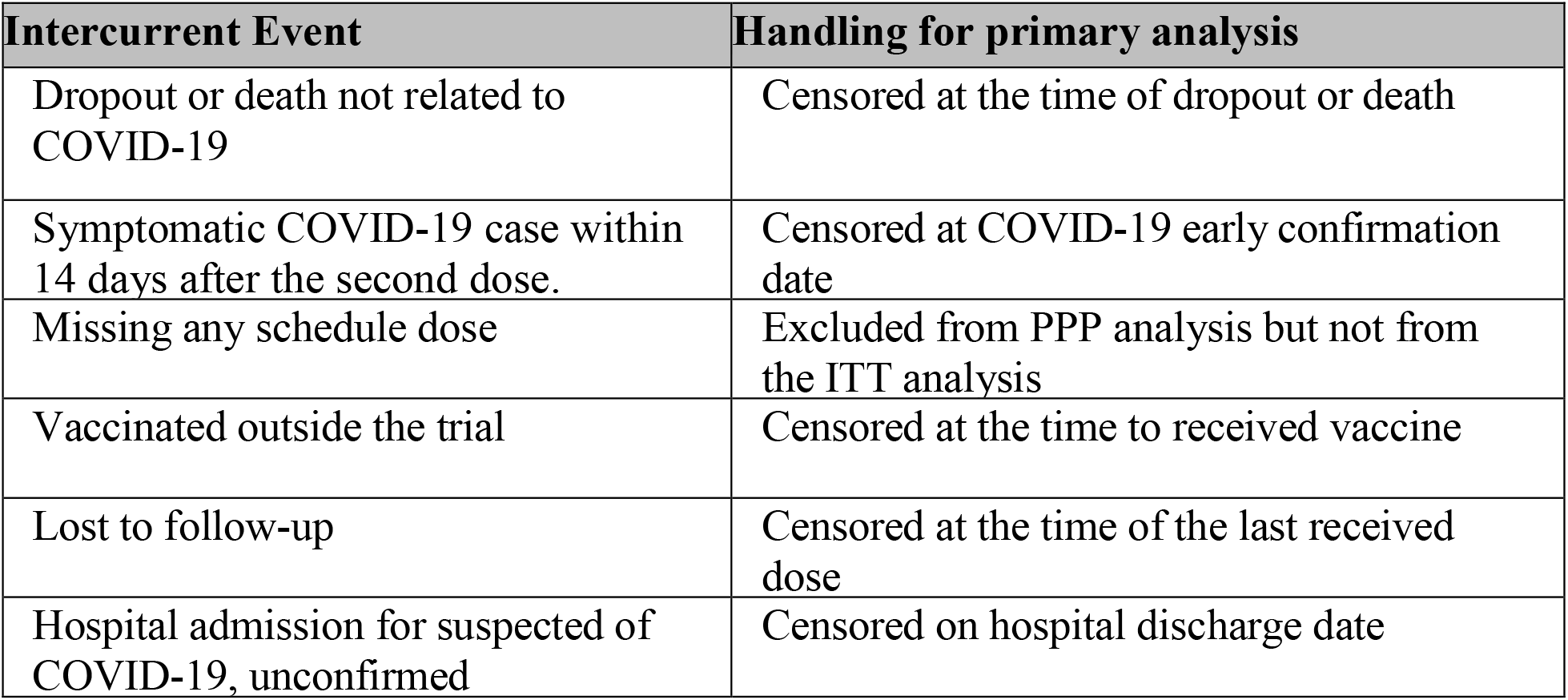

**S4. Figure S1.**
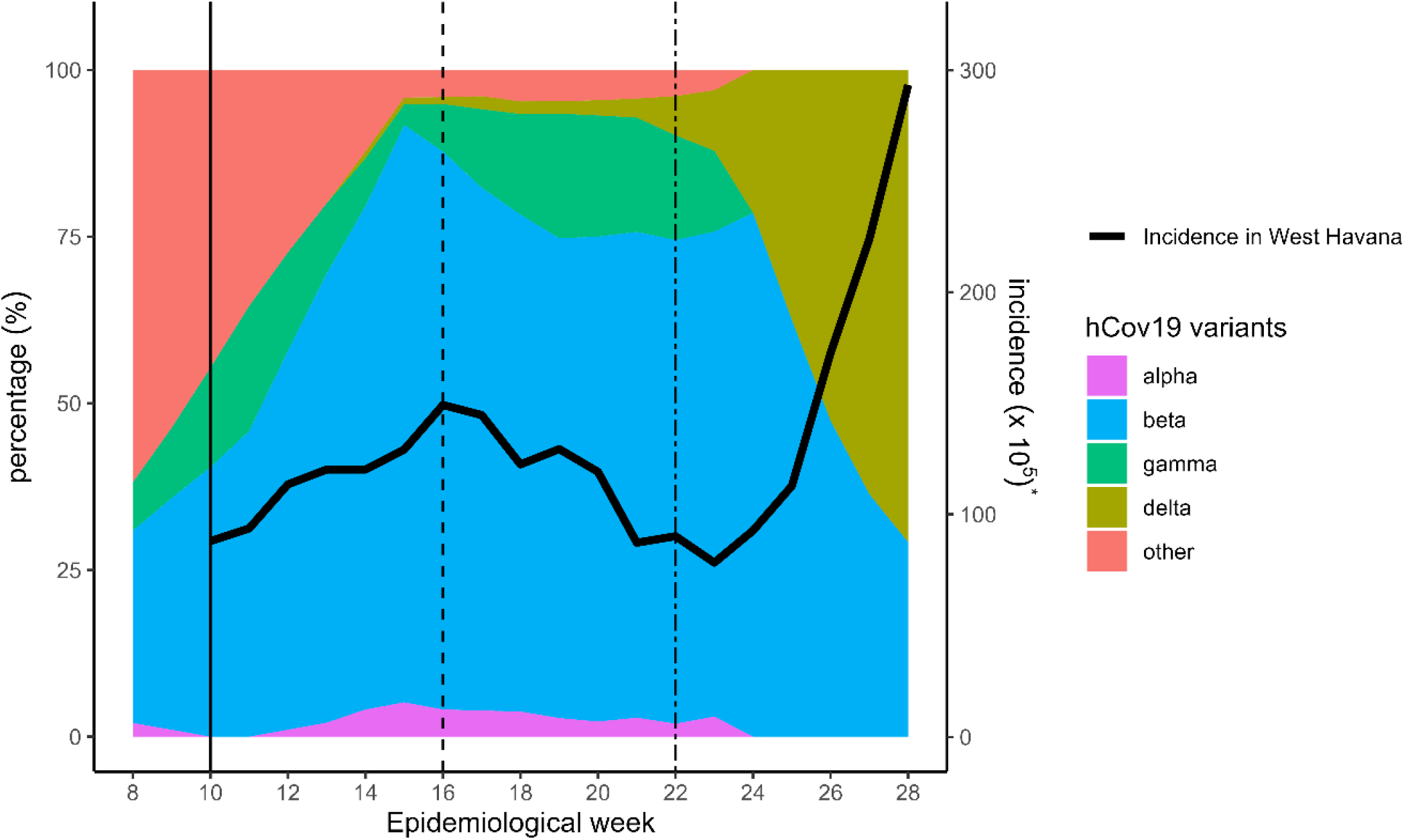
Relative frequency of circulating SARS-CoV-2 variants of concern and incidence rate of symptomatic infection in West Havana around the study period. ─ dose 1; -- start So2 efficacy evaluation; ─.─ start So2P efficacy evaluation * Incidence of symptomatic infection per 10^5^ in 19-80 years old in the population of the study area (8 municipalities of west Havana)

**S5. Table S3.**
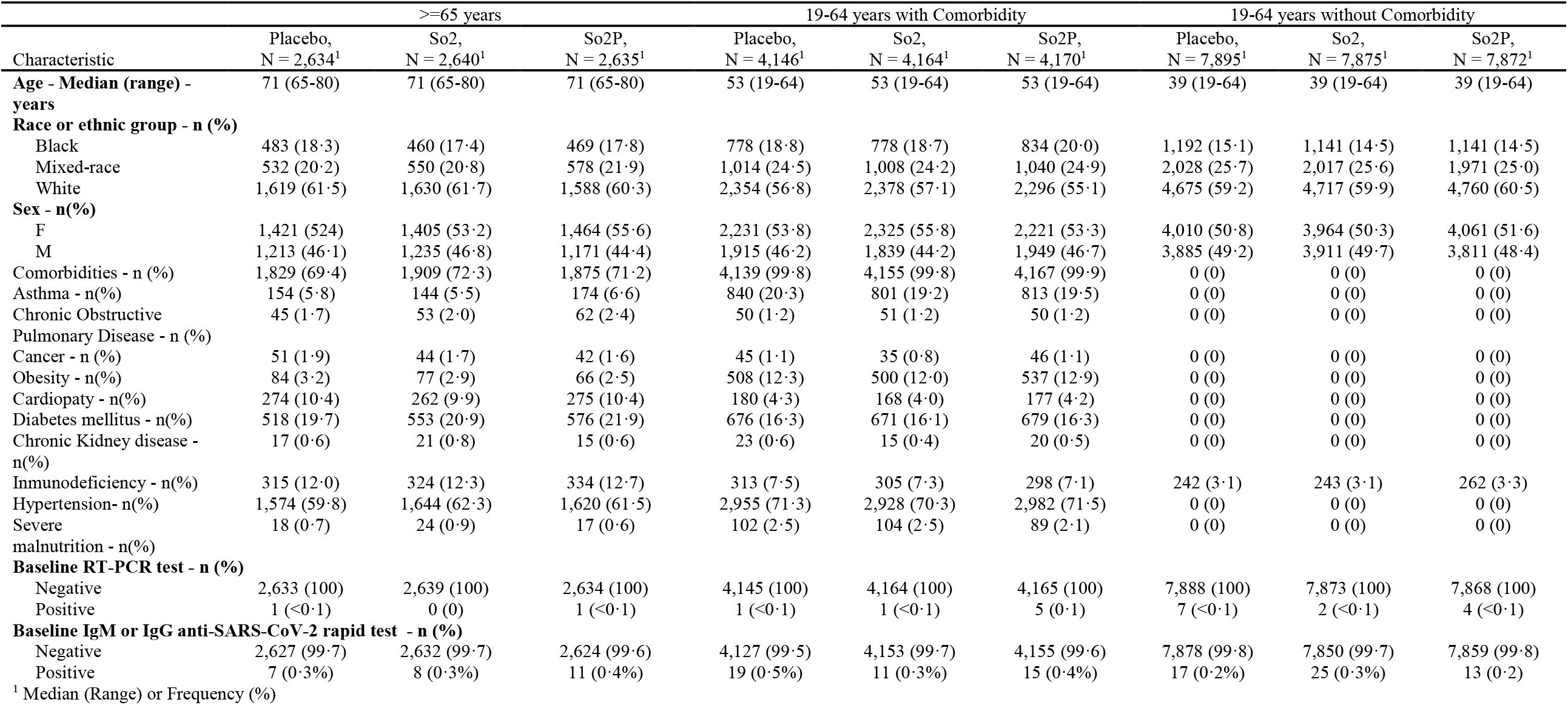
Baseline Demographics and Characteristics by Age and risk of severe COVID-19.

**S6. Table S4.** Solicited vaccine-associated Adverse Events (VAAEs) Within 7 Days After First Injection by Grade.

|  | Overall Safety Set |  |  | 19-64 years |  |  | ≥65 years |  |  |
| --- | --- | --- | --- | --- | --- | --- | --- | --- | --- |
|  | Placebo | So2 | So2P | Placebo | So2 | So2P | Placebo | So2 | So2P |
|  | N = 14,675 | N = 14,679 | N = 14,677 | N = 12,041 | N = 12,039 | N = 12,042 | N = 2,634 | N = 2,640 | N = 2,635 |
| Any solicited AR | 540 (3.7) | 1200 (8.2) | 1300 (8.9) | 442 (3.7) | 1053 (8.7) | 1125 (9.3) | 98 (3.7) | 147 (5.6) | 175 (6.6) |
| Grade 1 | 521 (3.5) | 1161 (7.9) | 1254 (8.5) | 427 (3.5) | 1015 (8.4) | 1084 (9.0) | 94 (3.6) | 146 (5.5) | 170 (6.4) |
| Grade 2 | 18 (0.1) | 37 (0.2) | 44 (0.3) | 15 (0.1) | 36 (0.3) | 39 (0.3) | 3 (0.1) | 1 (<0.1) | 5 (0.2) |
| Grade 3 | 1 (<0.1) | 2 (<0.1) | 2 (<0.1) | 0 | 2 (<0.1) | 2 (<0.1) | 1 (<0.1) | 0 | 0 |
| Any local AR | 382 (2.6) | 1079 (7.4) | 1178 (8.0) | 312 (2.6) | 950 (7.9) | 1027 (8.5) | 70 (2.5) | 129 (4.9) | 151 (5.7) |
| Grade 1 | 374 (2.5) | 1049 (7.1) | 1152 (7.8) | 305 (2.5) | 920 (7.6) | 1002 (8.3) | 69 (2.6) | 129 (4.9) | 150 (5.7) |
| Grade 2 | 8 (<0.1) | 28 (0.2) | 25 (0.2) | 7 (<0.1) | 28 (0.2) | 24 (0.2) | 1 (<0.1) | 0 | 1 (<0.1) |
| Grade 3 | 0 | 2 (<0.1) | 1 (<0.1) | 0 | 2 (<0.1) | 1 (<0.1) | 0 | 0 | 0 |
| Local AR |  |  |  |  |  |  |  |  |  |
| Pain | 347 (2.4) | 1029 (7.0) | 1096 (7.5) | 286 (2.4) | 907 (7.5) | 961 (8.0) | 61 (2.3) | 122 (4.6) | 135 (5.1) |
| Grade 1 | 340 (2.3) | 1000 (6.8) | 1072 (7.3) | 279 (2.3) | 878 (7.3) | 938 (7.8) | 61 (2.3) | 122 (4.6) | 134 (5.1) |
| Grade 2 | 7 (<0.1) | 27 (0.2) | 23 (0.2) | 7 (<0.1) | 27 (0.2) | 22 (0.2) | 0 | 0 | 1 (<0.1) |
| Grade 3 | 0 | 2 (<0.1) | 1 (<0.1) | 0 | 2 (<0.1) | 1 (<0.1) | 0 | 0 | 0 |
| Erythema | 16 (0.1) | 43 (0.3) | 64 (0.4) | 14 (0.1) | 37 (0.3) | 55 (0.5) | 2 (<0.1) | 6 (0.2) | 9 (0.3) |
| Grade 1 | 15 (0.1) | 42 (0.3) | 62 (0.4) | 14 (0.1) | 36 (0.3) | 53 (0.4) | 1 (<0.1) | 6 (0.2) | 9 (0.3) |
| Grade 2 | 1 (<0.1) | 1 (<0.1) | 2 (<0.1) | 0 | 1 (<0.1) | 2 (<0.1) | 1 (<0.1) | 0 | 0 |
| Grade 3 | 0 | 0 | 0 | 0 | 0 | 0 | 0 | 0 | 0 |
| Local Warmth | 25 (0.2) | 71 (0.5) | 67 (0.4) | 20 (0.2) | 63 (0.5) | 58 (0.5) | 5 (0.2) | 8 (0.3) | 9 (0.3) |
| Grade 1 | 25 (0.2) | 69 (0.5) | 66 (0.4) | 20 (0.2) | 61 (0.5) | 57 (0.5) | 5 (0.2) | 8 (0.3) | 9 (0.3) |
| Grade 2 | 0 | 2 (<0.1) | 1 (<0.1) | 0 | 2 (<0.1) | 1 (<0.1) | 0 | 0 | 0 |
| Grade 3 | 0 | 0 | 0 | 0 | 0 | 0 | 0 | 0 | 0 |
| Induration | 20 (0.1) | 91 (0.6) | 122 (0.8) | 16 (0.1) | 74 (0.6) | 108 (0.9) | 4 (0.2) | 17 (0.6) | 14 (0.5) |
| Grade 1 | 20 (0.1) | 88 (0.6) | 117 (0.8) | 16 (0.1) | 71 (0.6) | 103 (0.9) | 4 (0.2) | 17 (0.6) | 14 (0.5) |
| Grade 2 | 0 | 3 (<0.1) | 5 (<0.1) | 0 | 3 (<0.1) | 5 (<0.1) | 0 | 0 | 0 |
| Grade 3 | 0 | 0 | 0 | 0 | 0 | 0 | 0 | 0 | 0 |
| Swelling | 35 (0.2) | 127 (0.9) | 176 (1.2) | 30 (0.2) | 118 (1.0) | 149 (1.2) | 5 (0.2) | 9 (0.3) | 27 (1.0) |
| Grade 1 | 34 (0.2) | 124 (0.8) | 174 (1.2) | 29 (0.2) | 115 (0.9) | 148 (1.2) | 5 (0.2) | 9 (0.3) | 26 (1.0) |
| Grade 2 | 1 (<0.1) | 3 (<0.1) | 2 (<0.1) | 1 (<0.1) | 3 (<0.1) | 1 (<0.1) | 0 | 0 | 1 (<0.1) |
| Grade 3 | 0 | 0 | 0 | 0 | 0 | 0 | 0 | 0 | 0 |
| Any Systemic AR | 212 (1.4) | 247 (1.7) | 288 (2.0) | 177 (1.5) | 211 (1.8) | 238 (2.0) | 35 (1.3) | 36 (1.4) | 50 (1.9) |
| Grade 1 | 200 (1.4) | 234 (1.6) | 265 (1.8) | 168 (1.4) | 199 (1.6) | 220 (1.8) | 32 (1.2) | 35 (1.3) | 45 (1.7) |
| Grade 2 | 11 (<0.1) | 13 (<0.1) | 21 (0.1) | 9 (<0.1) | 12 (0.1) | 16 (0.1) | 2 (<0.1) | 1 (<0.1) | 5 (0.2) |
| Grade 3 | 1 (<0.1) | 0 | 2 (<0.1) | 0 | 0 | 2 (<0.1) | 1 (<0.1) | 0 | 0 |
| General discomfort | 172 (1.2) | 209 (1.4) | 239 (1.6) | 140 (1.2) | 176 (1.5) | 196 (1.6) | 32 (1.2) | 33 (1.2) | 43 (1.6) |
| Grade 1 | 163 (1.1) | 200 (1.4) | 221 (1.5) | 133 (1.1) | 168 (1.4) | 181 (1.5) | 30 (1.1) | 32 (1.2) | 40 (1.5) |
| Grade 2 | 8 (<0.1) | 9 (<0.1) | 16 (0.1) | 7 (<0.1) | 8 (<0.1) | 13 (0.1) | 1 (<0.1) | 1 (<0.1) | 3 (0.1) |
| Grade 3 | 1 (<0.1) | 0 | 2 (<0.1) | 0 | 0 | 2 (<0.1) | 1 (<0.1) | 0 | 0 |
| Rash | 6 (<0.1) | 2 (<0.1) | 9 (<0.1) | 5 (<0.1) | 2 (<0.1) | 8 (<0.1) | 1 (<0.1) | 0 | 1 (<0.1) |
| Grade 1 | 4 (<0.1) | 1 (<0.1) | 8 (<0.1) | 4 (<0.1) | 1 (<0.1) | 7 (<0.1) | 0 | 0 | 1 (<0.1) |
| Grade 2 | 2 (<0.1) | 1 (<0.1) | 1 (<0.1) | 1 (<0.1) | 1 (<0.1) | 1 (<0.1) | 1 (<0.1) | 0 | 0 |
| Grade 3 | 0 | 0 | 0 | 0 | 0 | 0 | 0 | 0 | 0 |
| Fever | 41 (0.3) | 45 (0.3) | 53 (0.4) | 39 (0.3) | 42 (0.4) | 46 (0.4) | 2 (<0.1) | 3 (0.1) | 7 (0.3) |
| Grade 1 | 40 (0.3) | 41 (0.3) | 47 (0.3) | 38 (0.3) | 38 (0.3) | 43 (0.4) | 2 (<0.1) | 3 (0.1) | 4 (0.2) |
| Grade 2 | 1 (<0.1) | 4 (<0.1) | 6 (<0.1) | 1 (<0.1) | 4 (<0.1) | 3 (<0.1) | 0 | 0 | 3 (0.1) |
| Grade 3 | 0 | 0 | 0 | 0 | 0 | 0 | 0 | 0 | 0 |

**S7. Table S5.** Solicited vaccine-associated Adverse Events (VAAEs) Within 7 Days After Second Injection by Grade.

|  | Overall Safety Set |  |  | 19-64 years |  |  | ≥65 years |  |  |
| --- | --- | --- | --- | --- | --- | --- | --- | --- | --- |
|  | Placebo | So2 | So2P | Placebo | So2 | So2P | Placebo | So2 | So2P |
|  | N = 14,400 | N = 14,447 | N = 14,421 | N = 11,801 | N = 11,841 | N = 11,832 | N = 2,599 | N = 2,606 | N = 2,589 |
| Any solicited AR | 78 (0.5) | 197 (1.4) | 469 (3.2) | 66 (0.6) | 171 (1.4) | 386 (3.3) | 12 (0.5) | 26 (1.0) | 83 (3.2) |
| Grade 1 | 73 (0.5) | 188 (1.3) | 445 (3.1) | 63 (0.5) | 162 (1.4) | 365 (3.1) | 10 (0.4) | 26 (1.0) | 80 (3.0) |
| Grade 2 | 5 (<0.1) | 8 (<0.1) | 22 (0.2) | 3 (<0.1) | 8 (0.1) | 19 (0.2) | 2 (<0.1) | 0 | 3 (0.1) |
| Grade 3 | 0 | 1 (<0.1) | 2 (<0.1) | 0 | 1 (<0.1) | 2 (<0.1) | 0 | 0 | 0 |
| Any local AR | 57 (0.4) | 162 (1.1) | 394 (2.7) | 47 (0.4) | 144 (1.2) | 327 (2.8) | 10 (0.4) | 18 (0.7) | 67 (2.6) |
| Grade 1 | 54 (0.4) | 157 (1.1) | 381 (2.6) | 45 (0.4) | 139 (1.2) | 316 (2.7) | 9 (0.3) | 18 (0.7) | 65 (2.5) |
| Grade 2 | 3 (<0.1) | 4 (<0.1) | 12 (<0.1) | 2 (<0.1) | 4 (<0.1) | 10 (<0.1) | 1 (<0.1) | 0 | 2 (<0.1) |
| Grade 3 | 0 | 1 (<0.1) | 1 (<0.1) | 0 | 1 (<0.1) | 1 (<0.1) | 0 | 0 | 0 |
| Local AR |  |  |  |  |  |  |  |  |  |
| Pain | 50 (0.4) | 147 (1.0) | 367 (2.5) | 40 (0.3) | 130 (1.1) | 305 (2.6) | 10 (0.4) | 17 (0.6) | 62 (2.3) |
| Grade 1 | 47 (0.3) | 142 (1.0) | 355 (2.4) | 38 (0.3) | 125 (1.1) | 294 (2.5) | 9 (0.4) | 17 (0.6) | 61 (2.3) |
| Grade 2 | 3 (<0.1) | 4 (<0.1) | 11 (<0.1) | 2 (<0.1) | 4 (<0.1) | 10 (<0.1) | 1 (<0.1) | 0 | 1 (<0.1) |
| Grade 3 | 0 | 1 (<0.1) | 1 (<0.1) | 0 | 1 (<0.1) | 1 (<0.1) | 0 | 0 | 0 |
| Erythema | 0 | 10 (<0.1) | 25 (0.2) | 0 | 9 (<0.1) | 17 (0.1) | 0 | 1 (<0.1) | 8 (0.3) |
| Grade 1 | 0 | 10 (<0.1) | 25 (0.2) | 0 | 9 (<0.1) | 17 (0.1) | 0 | 1 (<0.1) | 8 (0.3) |
| Grade 2 | 0 | 0 | 0 | 0 | 0 | 0 | 0 | 0 | 0 |
| Grade 3 | 0 | 0 | 0 | 0 | 0 | 0 | 0 | 0 | 0 |
| Local Warmth | 5 (<0.1) | 14 (0.1) | 22 (0.2) | 4 (<0.1) | 11 (<0.1) | 18 (0.2) | 1 (<0.1) | 3 (0.1) | 4 (0.2) |
| Grade 1 | 5 (<0.1) | 14 (0.1) | 22 (0.2) | 4 (<0.1) | 11 (<0.1) | 18 (0.2) | 1 (<0.1) | 3 (0.1) | 4 (0.2) |
| Grade 2 | 0 | 0 | 0 | 0 | 0 | 0 | 0 | 0 | 0 |
| Grade 3 | 0 | 0 | 0 | 0 | 0 | 0 | 0 | 0 | 0 |
| Induration | 6 (<0.1) | 14 (0.1) | 39 (0.3) | 4 (<0.1) | 12 (0.1) | 31 (0.3) | 2 (<0.1) | 2 (<0.1) | 8 (0.3) |
| Grade 1 | 6 (<0.1) | 14 (0.1) | 38 (0.3) | 4 (<0.1) | 12 (0.1) | 30 (0.2) | 2 (<0.1) | 2 (<0.1) | 8 (0.3) |
| Grade 2 | 0 | 0 | 1 (<0.1) | 0 | 0 | 1 (<0.1) | 0 | 0 | 0 |
| Grade 3 | 0 | 0 | 0 | 0 | 0 | 0 | 0 | 0 | 0 |
| Swelling | 12 (<0.1) | 32 (0.2) | 82 (0.6) | 12 (0.1) | 30 (0.2) | 70 (0.6) | 0 | 2 (<0.1) | 12 (0.5) |
| Grade 1 | 12 (<0.1) | 32 (0.2) | 79 (0.6) | 12 (0.1) | 30 (0.2) | 68 (0.6) | 0 | 2 (<0.1) | 11 (0.4) |
| Grade 2 | 0 | 0 | 3 (<0.1) | 0 | 0 | 2 (<0.1) | 0 | 0 | 1 (<0.1) |
| Grade 3 | 0 | 0 | 0 | 0 | 0 | 0 | 0 | 0 | 0 |
| Any Systemic AR | 27 (0.2) | 66 (0.4) | 141 (1.0) | 23 (0.2) | 56 (0.5) | 111 (0.9) | 4 (0.2) | 10 (0.4) | 30 (1.2) |
| Grade 1 | 25 (0.2) | 61 (0.4) | 129 (0.9) | 22 (0.2) | 51 (0.4) | 100 (0.8) | 3 (0.1) | 10 (0.4) | 29 (1.1) |
| Grade 2 | 2 (<0.1) | 5 (<0.1) | 11 (<0.1) | 1 (<0.1) | 5 (<0.1) | 10 (<0.1) | 1 (<0.1) | 0 | 1 (<0.1) |
| Grade 3 | 0 | 0 | 1 (<0.1) | 0 | 0 | 1 (<0.1) | 0 | 0 | 0 |
| General discomfort | 21 (0.2) | 57 (0.4) | 121 (0.8) | 17 (0.1) | 48 (0.4) | 95 (0.8) | 4 (0.2) | 9 (0.4) | 26 (0.2) |
| Grade 1 | 20 (0.0) | 53 (0.4) | 111 (0.8) | 17 (0.1) | 44 (0.4) | 86 (0.7) | 3 (0.1) | 9 (0.4) | 25 (0.2) |
| Grade 2 | 1 (<0.1) | 4 (<0.1) | 9 (<0.1) | 0 | 4 (<0.1) | 8 (<0.1) | 1 (<0.1) | 0 | 1 (<0.1) |
| Grade 3 | 0 | 0 | 1 (<0.1) | 0 | 0 | 1 (<0.1) | 0 | 0 | 0 |
| Rash | 2 (<0.1) | 0 | 3 (<0.1) | 2 (<0.1) | 0 | 2 (<0.1) | 0 | 0 | 1 (<0.1) |
| Grade 1 | 2 (<0.1) | 0 | 3 (<0.1) | 2 (<0.1) | 0 | 2 (<0.1) | 0 | 0 | 1 (<0.1) |
| Grade 2 | 0 | 0 | 0 | 0 | 0 | 0 | 0 | 0 | 0 |
| Grade 3 | 0 | 0 | 0 | 0 | 0 | 0 | 0 | 0 | 0 |
| Fever | 5 (<0.1) | 15 (0.1) | 28 (0.2) | 5 (<0.1) | 14 (0.1) | 21 (0.2) | 0 | 1 (<0.1) | 7 (0.3) |
| Grade 1 | 4 (<0.1) | 13 (<0.1) | 25 (0.2) | 4 (<0.1) | 12 (0.1) | 19 (0.2) | 0 | 1 (<0.1) | 6 (0.2) |
| Grade 2 | 1 (<0.1) | 2 (<0.1) | 3 (<0.1) | 1 (<0.1) | 2 (<0.1) | 2 (<0.1) | 0 | 0 | 1 (<0.1) |
| Grade 3 | 0 | 0 | 0 | 0 | 0 | 0 | 0 | 0 | 0 |

**S8. Table S6.** Solicited vaccine-associated Adverse Events (VAAEs) Within 7 Days After Third Injection by Grade.

|  |  | Overall Safety Set | 19-64 years | ≥65 years |
| --- | --- | --- | --- | --- |
|  |  | So2P<br>N = 13,979 | So2P<br>N = 11,475 | So2P<br>N = 2,504 |
| Any solicited AR |  | 41 (0.3) | 35 (0.3) | 6 (0.2) |
|  | Grade 1 | 40 (0.3) | 34 (0.3) | 6 (0.2) |
|  | Grade 2 | 1 (<0.1) | 1 (<0.1) | 0 |
|  | Grade 3 | 0 | 0 | 0 |
| Any local AR |  | 37 (0.3) | 32 (0.3) | 5 (0.2) |
|  | Grade 1 | 36 (0.3) | 31 (0.3) | 5 (0.2) |
|  | Grade 2 | 1 (<0.1) | 1 (<0.1) | 0 |
|  | Grade 3 | 0 | 0 | 0 |
| Local AR |  |  |  |  |
| Pain |  | 25 (0.2) | 21 (0.2) | 4 (0.2) |
|  | Grade 1 | 25 (0.2) | 21 (0.2) | 4 (0.2) |
|  | Grade 2 | 0 | 0 | 0 |
|  | Grade 3 | 0 | 0 | 0 |
| Erythema |  | 7 (<0.1) | 7 (<0.1) | 0 |
|  | Grade 1 | 7 (<0.1) | 7 (<0.1) | 0 |
|  | Grade 2 | 0 | 0 | 0 |
|  | Grade 3 | 0 | 0 | 0 |
| Local Warmth |  | 6 (<0.1) | 6 (<0.1) | 0 |
|  | Grade 1 | 6 (<0.1) | 6 (<0.1) | 0 |
|  | Grade 2 | 0 | 0 | 0 |
|  | Grade 3 | 0 | 0 | 0 |
| Induration |  | 4 (<0.1) | 4 (<0.1) | 0 |
|  | Grade 1 | 4 (<0.1) | 4 (<0.1) | 0 |
|  | Grade 2 | 0 | 0 | 0 |
|  | Grade 3 | 0 | 0 | 0 |
| Swelling |  | 14 (0.1) | 13 (0.1) | 1 (<0.1) |
|  | Grade 1 | 13 (<0.1) | 12 (0.1) | 1 (<0.1) |
|  | Grade 2 | 1 (<0.1) | 1 (<0.1) | 0 |
|  | Grade 3 | 0 | 0 | 0 |
| Any Systemic AR |  | 8 (<0.1) | 6 (<0.1) | 2 (<0.1) |
|  | Grade 1 | 8 (<0.1) | 6 (<0.1) | 2 (<0.1) |
|  | Grade 2 | 0 | 0 | 0 |
|  | Grade 3 | 0 | 0 | 0 |
| General discomfort |  | 4 (<0.1) | 3 (<0.1) | 1 (<0.1) |
|  | Grade 1 | 4 (<0.1) | 3 (<0.1) | 1 (<0.1) |
|  | Grade 2 | 0 | 0 | 0 |
|  | Grade 3 | 0 | 0 | 0 |
| Rash |  | 3 (<0.1) | 2 (<0.1) | 1 (<0.1) |
|  | Grade 1 | 3 (<0.1) | 2 (<0.1) | 1 (<0.1) |
|  | Grade 2 | 0 | 0 | 0 |
|  | Grade 3 | 0 | 0 | 0 |
| Fever |  | 1 (<0.1) | 1 (<0.1) | 0 |
|  | Grade 1 | 1 (<0.1) | 1 (<0.1) | 0 |
|  | Grade 2 | 0 | 0 | 0 |
|  | Grade 3 | 0 | 0 | 0 |

**S9. Table S7.**
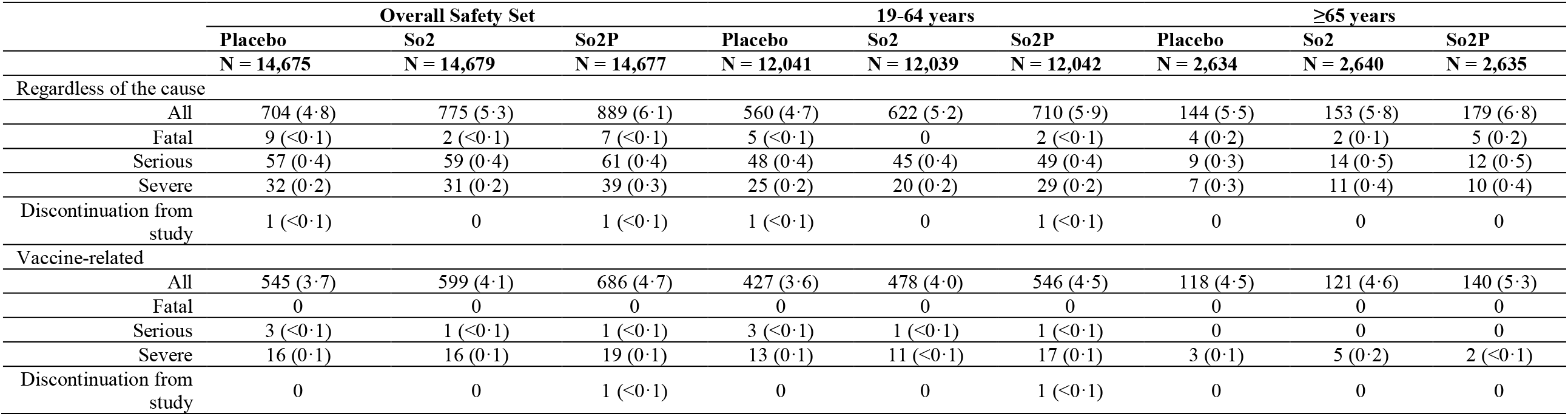
Unsolicited AEs Within 28 Days After Any Injection, Overall Safety Set.

|  | Overall Safety Set |  |  | 19-64 years |  |  | ≥65 years |  |  |
| --- | --- | --- | --- | --- | --- | --- | --- | --- | --- |
|  | Placebo<br>N = 14,675 | So2<br>N = 14,679 | So2P<br>N = 14,677 | Placebo<br>N = 12,041 | So2<br>N = 12,039 | So2P<br>N = 12,042 | Placebo<br>N = 2,634 | So2<br>N = 2,640 | So2P<br>N = 2,635 |
| Regardless of the cause |  |  |  |  |  |  |  |  |  |
| All | 704 (4.8) | 775 (5.3) | 889 (6.1) | 560 (4.7) | 622 (5.2) | 710 (5.9) | 144 (5.5) | 153 (5.8) | 179 (6.8) |
| Fatal | 9 (<0.1) | 2 (<0.1) | 7 (<0.1) | 5 (<0.1) | 0 | 2 (<0.1) | 4 (0.2) | 2 (0.1) | 5 (0.2) |
| Serious | 57 (0.4) | 59 (0.4) | 61 (0.4) | 48 (0.4) | 45 (0.4) | 49 (0.4) | 9 (0.3) | 14 (0.5) | 12 (0.5) |
| Severe | 32 (0.2) | 31 (0.2) | 39 (0.3) | 25 (0.2) | 20 (0.2) | 29 (0.2) | 7 (0.3) | 11 (0.4) | 10 (0.4) |
| Discontinuation from study | 1 (<0.1) | 0 | 1 (<0.1) | 1 (<0.1) | 0 | 1 (<0.1) | 0 | 0 | 0 |
| Vaccine-related |  |  |  |  |  |  |  |  |  |
| All | 545 (3.7) | 599 (4.1) | 686 (4.7) | 427 (3.6) | 478 (4.0) | 546 (4.5) | 118 (4.5) | 121 (4.6) | 140 (5.3) |
| Fatal | 0 | 0 | 0 | 0 | 0 | 0 | 0 | 0 | 0 |
| Serious | 3 (<0.1) | 1 (<0.1) | 1 (<0.1) | 3 (<0.1) | 1 (<0.1) | 1 (<0.1) | 0 | 0 | 0 |
| Severe | 16 (0.1) | 16 (0.1) | 19 (0.1) | 13 (0.1) | 11 (<0.1) | 17 (0.1) | 3 (0.1) | 5 (0.2) | 2 (<0.1) |
| Discontinuation from study | 0 | 0 | 1 (<0.1) | 0 | 0 | 1 (<0.1) | 0 | 0 | 0 |

**S10. Table S8.** Summary of Unsolicited Adverse Events (AEs) ≥0.1%, within 28 Days After Any Injection Overall Safety Set.

|  | **Placebo** | **So2** | **So2P** |
| --- | --- | --- | --- |
|  | **N = 14,675** | **N = 14,679** | **N = 14,677** |
| Number of participants reporting unsolicited AEs | 704 (4.8) | 775 (5.3) | 889 (6.1) |
| Number of unsolicited AEs | 851 | 931 | 1074 |
| Nervous system disorders |  |  |  |
| Headache | 143 (1.0) | 171 (1.2) | 214 (1.5) |
| Dizziness | 12 (<0.1) | 14 (0.1) | 8 (<0.1) |
| Somnolence | 12 (<0.1) | 9 (<0.1) | 14 (0.1) |
| Gastrointestinal disorders |  |  |  |
| Diarrhea | 19 (0.1) | 23 (0.2) | 36 (0.2) |
| Infections and infestations |  |  |  |
| Upper Acute respiratory infection | 87 (0.6) | 78 (0.5) | 87 (0.6) |
| Vascular disorders |  |  |  |
| High blood pressure measurement | 262 (1.8) | 314 (2.1) | 327 (2.2) |
| Musculoskeletal and connective tissue disorders |  |  |  |
| Myalgia | 8 (<0.1) | 16 (0.1) | 21 (0.1) |
| General disorders and administration site conditions |  |  |  |
| Mild fever | 13 (<0.1) | 19 (0.1) | 18 (0.1) |

**S11. Table S9.**
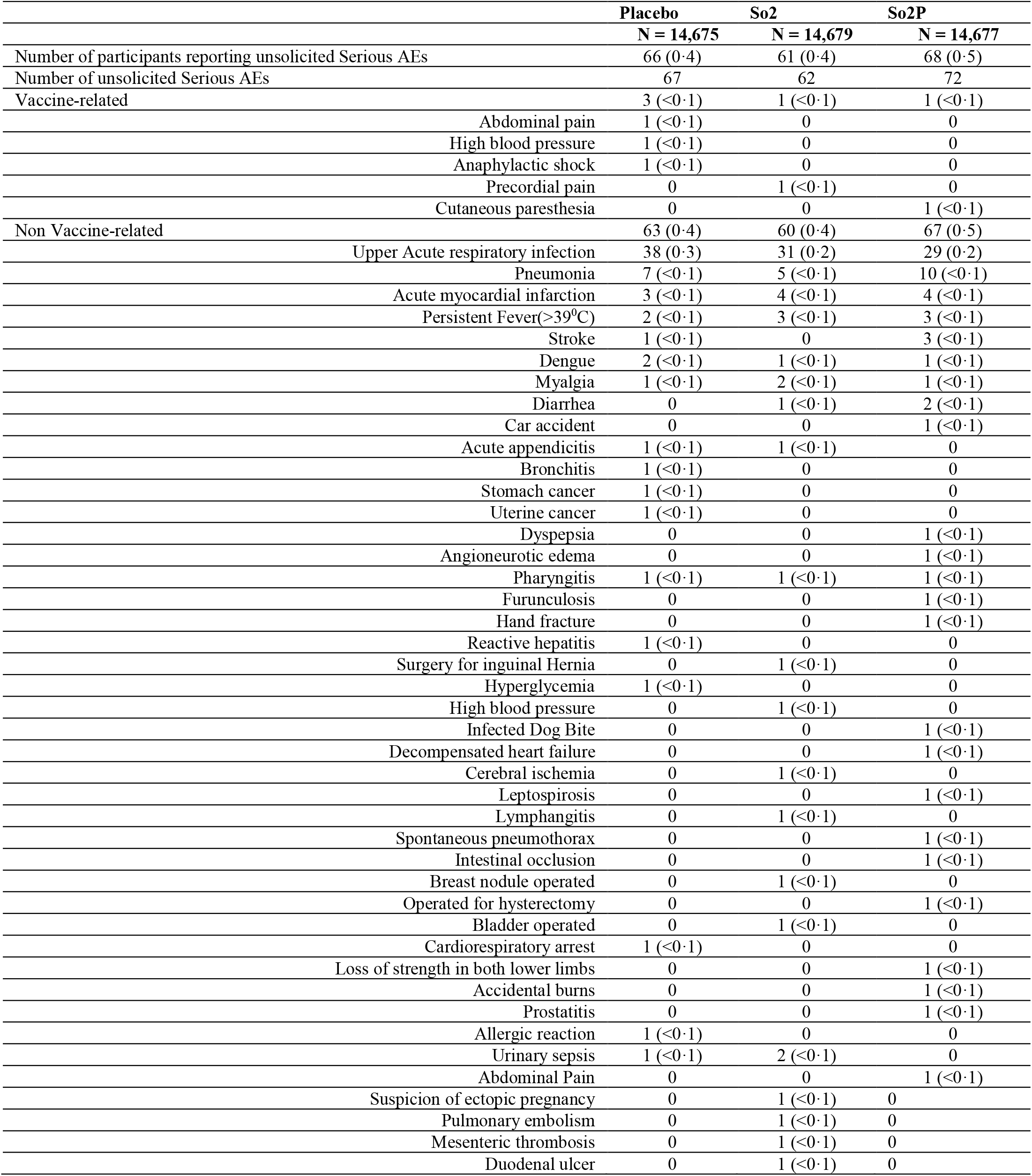
Summary of Unsolicited Serious Adverse Events (AEs), 28 Days After Any Injection Overall Safety Set.

**S12. Table S10.** Vaccine Efficacy for Prevention of severe COVID-19 disease in Per-Protocol Population (PPP).

|  | Two-dose So2 VE |  | So2P VE |  |
| --- | --- | --- | --- | --- |
| Secondary Endpoint | Placebo | So2 | Placebo | So2P |
| Severe Covid19 disease |  |  |  |  |
| n of events, PPP | 23 | 5 | 6 | 0 |
| Person- weeks | 70849.7 | 64890.0 | 19937.6 | 47070.4 |
| Incidence rate per 1,000 Person-Years (95% CI) | 16.9 (10.7; 25.4) | 4.0 (1.3; 9.4) | 15.7 (5.8; 34.2) | 0 (0; 4.1 ) |
| VE based on HR (95% CI)* |  | 74.9 (33.7; 90.5) | -- | 100.0 (-Inf; 100.0) |
| CI: Confidence Interval, VE: Vaccine Efficacy , defined as 1 - minus the hazard ratio (HR, Vaccine vs. placebo), and the 95% confidence interval was estimated with the use of a stratified Cox proportional hazards model |  |  |  |  |

**S13. Figure S2.**
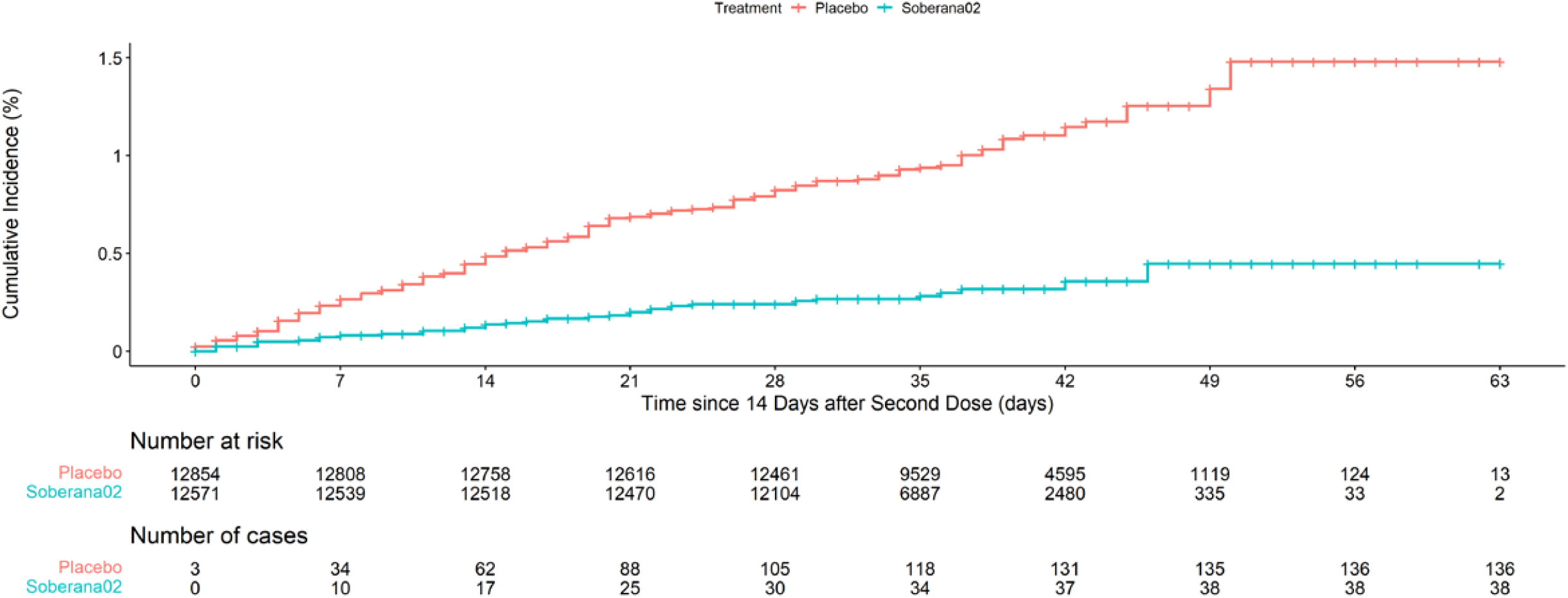
Cumulative incidence of Covid-19 for two doses of Soberana-02 and Placebo. Per-Protocol Analysis.

**S14. Figure S3.**
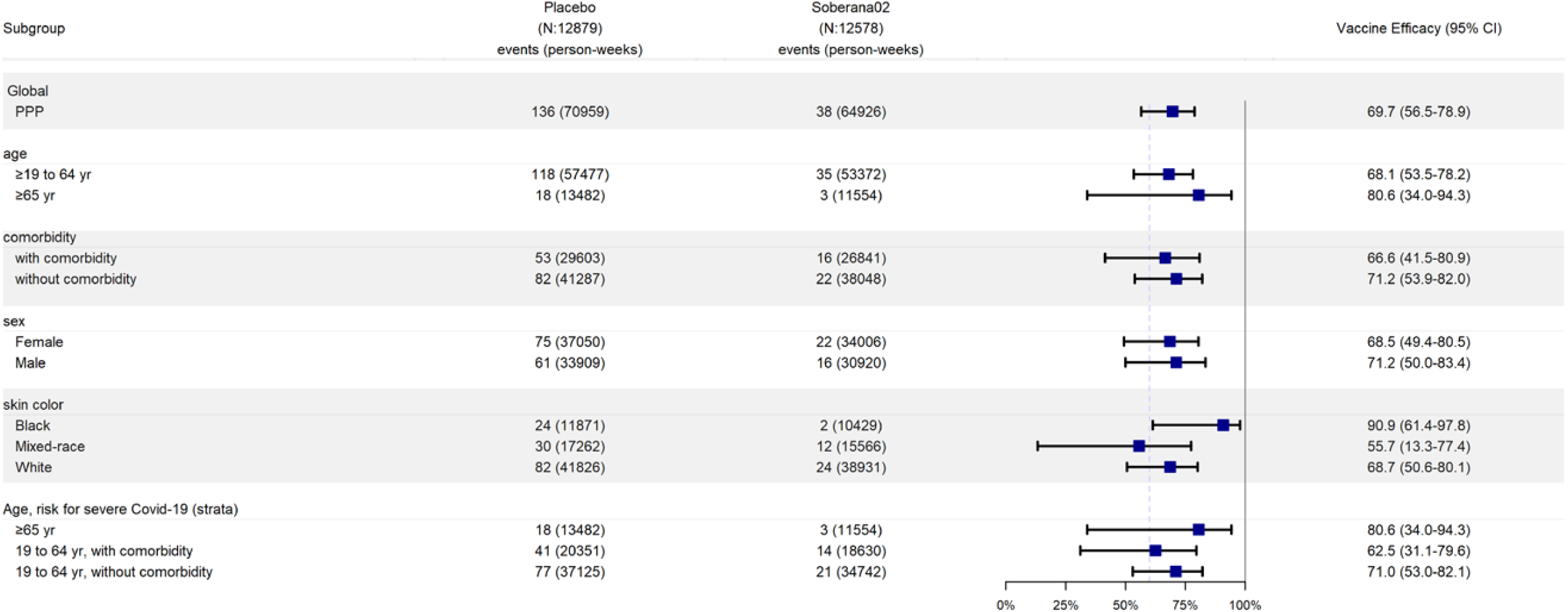
Vaccine Efficacy of two dose of Soberana-02 to Prevent Covid-19 disease in Subgroups, 14 days after 2^nd^ dose. The efficacy in preventing COVID-19 in various subgroups in the per-protocol population was based on adjudicated assessments starting 14 days after the second injection. Vaccine efficacy, defined as 1 minus the hazard ratio (So2 vs. placebo), and 95% confidence intervals were estimated with the use of a stratified Cox proportional hazards model.

**S15. Figure S4.**
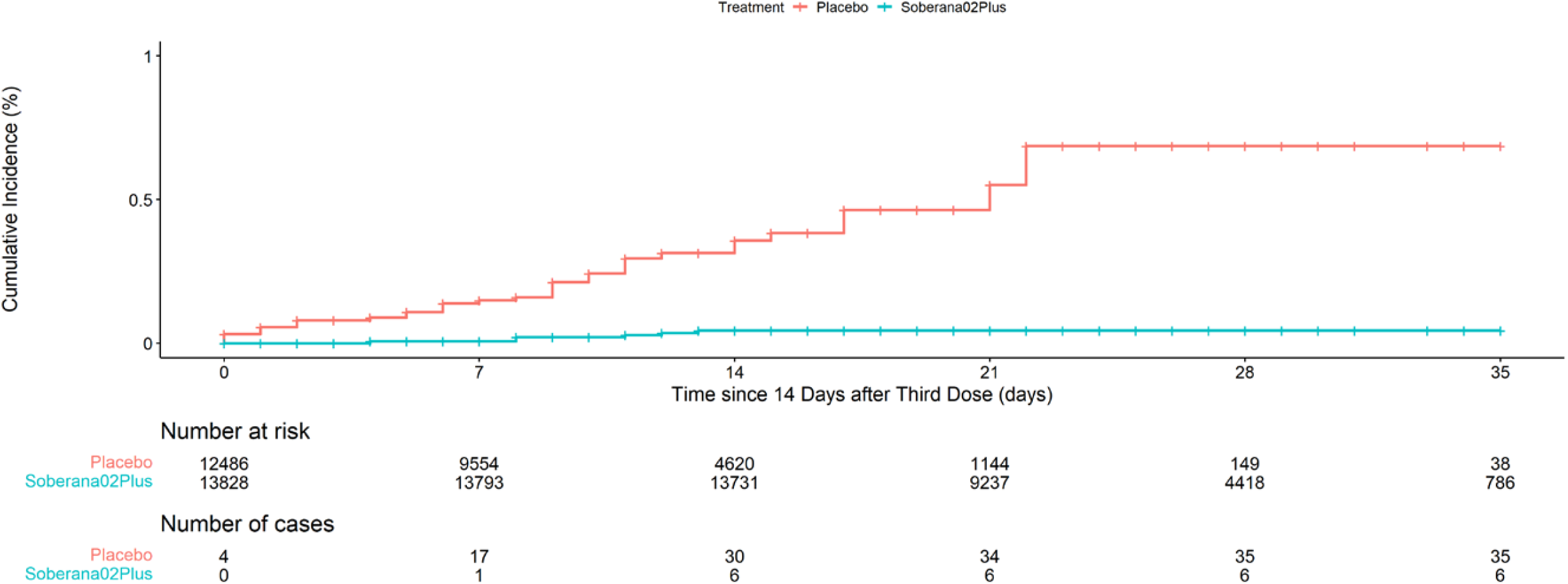
Cumulative incidence of Covid-19 for Soberana-02+Plus and Placebo. Per-Protocol Analysis.

**S16. Figure S5.**
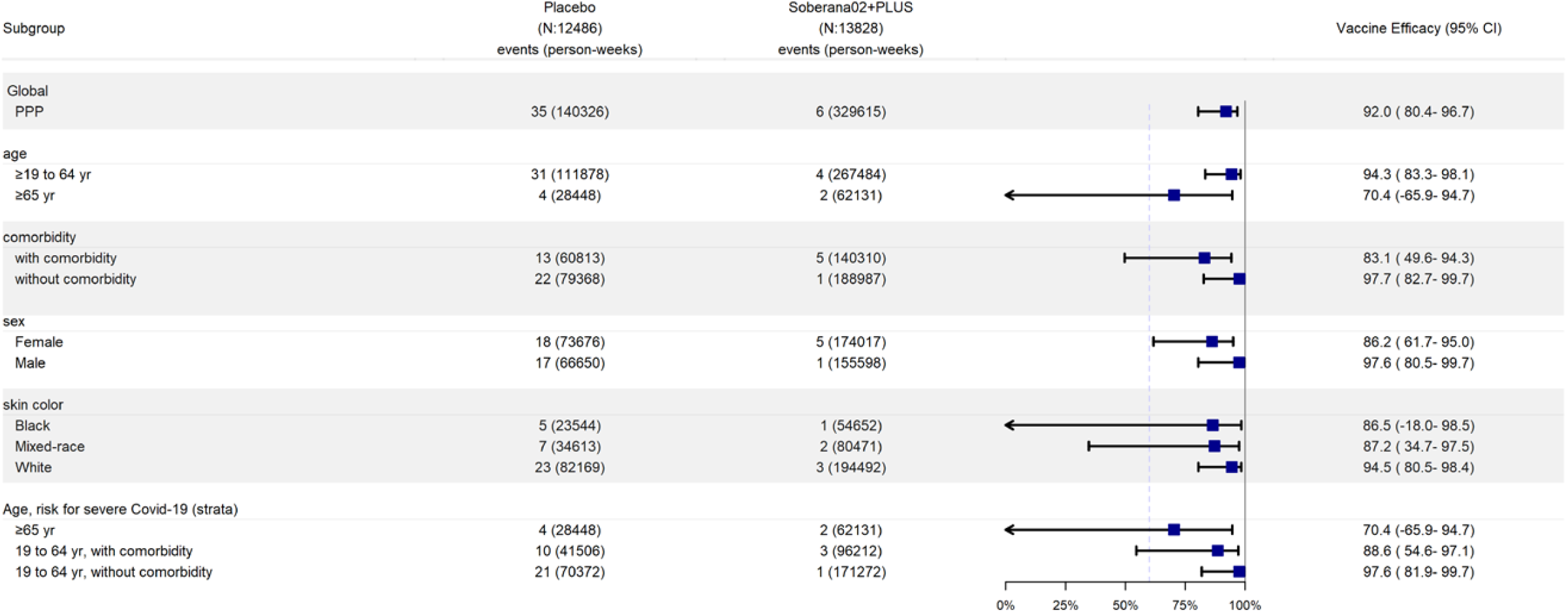
Vaccine Efficacy of heterologous schedule to Prevent Covid-19 disease in Subgroups 14 days after 3^rd^ dose. The efficacy in preventing COVID-19 in various subgroups in the per-protocol population was based on adjudicated assessments starting 14 days after the third injection (42 days after second injection for Placebo group). Vaccine efficacy, defined as 1 minus the hazard ratio (So2p vs. placebo), and 95% confidence intervals were estimated with the use of a stratified Cox proportional hazards model.

